# Improving vaccination coverage and offering vaccine to all school-age children will allow uninterrupted in-person schooling in King County, WA: Modeling analysis

**DOI:** 10.1101/2021.10.01.21264426

**Authors:** Chloe Bracis, Mia Moore, David A. Swan, Laura Matrajt, Larissa Anderson, Daniel B. Reeves, Eileen Burns, Joshua T. Schiffer, Dobromir Dimitrov

## Abstract

**Background:** The mass rollout of COVID vaccination in early 2021 allowed local and state authorities to relax mobility and social interaction regulations in spring 2021 including lifting all restrictions for vaccinated people and restoring in-person schooling. However, the emergence and rapid spread of highly transmissible variants combined with slowing down the pace of vaccination created uncertainty around the future trajectory of the epidemic. In this study we analyze the expected benefits of offering vaccination to children age 5-11 under differing conditions for in-person schooling.

**Methods:** We adapted a mathematical model of SARS-CoV-2 transmission, calibrated to data from King County, Washington, to handle multiple variants with increased transmissibility and virulence as well as differential vaccine efficacies against each variant. Reactive social distancing is implemented driven by fluctuations in the number of hospitalizations in the county. We simulate scenarios offering vaccination to children aged 5-11 with different starting dates and different proportions of physical interactions (PPI) in schools being restored. The impact of improving overall vaccination coverage among the eligible population is also explored. Cumulative hospitalizations, percentage reduction of hospitalizations and proportion of time at maximum social distancing over the 2021-2022 school year are reported.

**Findings:** In the base-case scenario with 85% vaccination coverage of 12+ year-olds, our model projects 4945 (median, IQR 4622-5341) total COVID-19 hospitalizations and 325 (median, IQR 264-400) pediatric hospitalizations if physical contacts at schools are fully restored (100% PPI) for the entire school year compared to 3675 (median, IQR 2311-4725) and 163 (median, IQR 95-226) if schools remained closed. Reducing contacts in schools to 75% PPI or 50% PPI through masking, ventilation and distancing is expected to decrease the overall cumulative hospitalizations by 2% and 4% respectively and youth hospitalizations by 8% and 23% respectively. Offering early vaccination to children aged 5-11 with 75% PPI is expected to prevent 756 (median, IQR 301-1434) hospitalizations and cut hospitalizations in the youngest age group in half compared to no vaccination. It will largely reduce the need of additional social distancing over the school year. If, in addition, 90% overall vaccination coverage is reached, 60% of remaining hospitalizations will be averted and the need of extra mitigation measures almost certainly avoided.

**Conclusions:** Our work highlights that in-person schooling is possible if reasonable precaution measures are taken at schools to reduced infectious contacts. Rapid vaccination of all school-aged children will provide meaningful reduction of the COVID health burden over this school year but only if implemented early. Finally, it remains critical to vaccinate as many people as possible to limit the morbidity and mortality associated with the current surge in Delta variant cases.

## Introduction

Several effective SARS-CoV-2 vaccines were initially approved for adults in early 2021 and later expanded to 12-16 year-old children in May, 2021, creating hope that the COVID pandemic could be controlled and that social distancing and mask mandates could be permanently relaxed. With a rapidly increasing proportion of the population vaccinated, local and state authorities began loosening COVID-19 restrictions in spring 2021 and lifting all restrictions for vaccinated people during summer of 2021 in many jurisdictions [1]

Concurrently, several highly transmissible variants of concern began to emerge. The B.1.1.7 (Alpha) variant quicky predominated in the first 3-4 months of the year but was soon replaced by the B.1.167.2 (Delta) variant over the summer[2,3]. Being approximately 60% more transmissible than the Alpha variant, the Delta variant has a greater proportional likelihood of hospitalization and need for ICU care [4],[5],[6].While vaccine efficacy data against these variants is still emerging, initial evidence suggests a decrease in protection against virologic proven symptomatic infection over 6 months since vaccination, while protection against severe disease and hospitalization is retained. [7] Together, the shifting dynamics created by ongoing vaccination, increased background immunity due to the surge of infections associated with the Delta variant, shifting characteristics of emerging viral variants and changes in social distancing and masking practices have created uncertainty around the future trajectory of the U.S. epidemic.

A critical unknown question is how to safely open schools and to estimate the impact of vaccinating children under the age of 12, particularly as recent clinical trials have reported high vaccine efficacy in children 5-11[8]. We have previously modeled the King County, Washington epidemic to estimate the impact of various policy decisions with the goal of drawing relevant conclusions for similar communities across the country. King County coordinated a rapid vaccination rollout which resulted in more than 76% of the eligible (12+ year) population fully vaccinated by August 15, 2021 [9]. As a large proportion of the unvaccinated population are school aged children, the question of the possible impact of reopening schools in fall 2021 in the context of a surging Delta variant is of critical importance. Schools in the county have been predominately closed since the beginning of the epidemic in March 2020 with most education being delivered online. Hybrid options of schooling were implemented throughout Washington state in spring 2021 without evident impact on the ongoing transmission [10]. All school districts in King County have returned to full-time in-person instruction as of September 2021, with only a small pilot online program for high-risk students. All districts developed guidelines to limit SARS-Cov-2 spread, which include the mandatory use of masks, adequate building ventilation, and other interventions to reduce contacts in schools below their pre-COVID levels. With school-age children less than 12 years old not eligible for vaccination and the high transmissibility of the dominant Delta variant, added school transmission may turn into a decisive factor for future epidemic outbreaks among the greater King County population.

In an earlier analysis, conducted during the initial COVID outbreak in spring, 2020, we concluded that school reopening would not significantly increase SARS-Cov-2 transmission if a well-coordinated combination of non-pharmaceutical mitigation measures were in place[11]. Other modeling studies suggest that infection rates through late November will exceed 75% and 90% among high school and elementary students respectively in the absence of masking and testing but that infection rates can be lowered significantly with both of these interventions[12]. In this study we use mathematical models to investigate the contribution that in-person schooling is expected to have on COVID hospitalizations, under the current projections of vaccination coverage in King County and the expansion of the Delta variant in the area. We also analyze the expected benefits of offering vaccination to children age 5-12 starting on Oct.1, 2021 relative to: 1) delaying this access by another 3 months or 2) continuing to vaccinate individuals 12 years and older only. Finally, we study how waning immunity of vaccine protection may affect these results and if it may change the recommendations regarding how schools should operate in King County.

## Methods

We adapted a previously developed deterministic compartment model which describes the epidemic dynamics in King County. [11,13–15] Our model (**Fig 1A** and **Fig S1**) stratifies the population by age (0-19 years, 20-49 years, 50-69 years, and 70+ years), infection status (susceptible, exposed, asymptomatic, pre-symptomatic, symptomatic mild, symptomatic severe), clinical status (undiagnosed, diagnosed, hospitalized), immunity (susceptible, vaccinated, recovered from natural infection), and infecting strain (original, Alpha, Delta).

**Figure 1.**
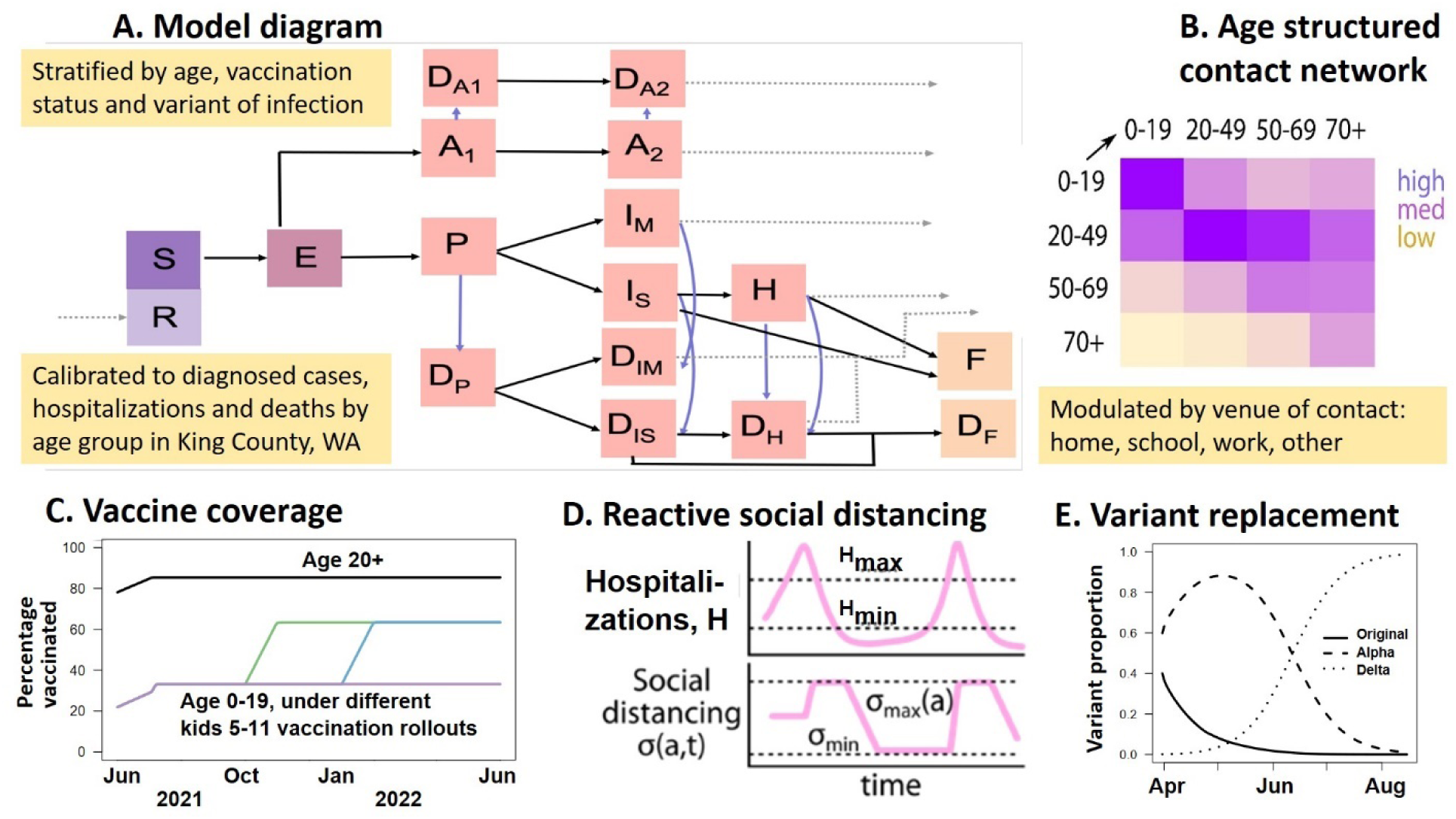
Modeling framework. **A)** Model diagram showing the progression of susceptible (S) individuals to exposed (E) who are not infectious, asymptomatic (A), pre-symptomatic (P), mild (I_M_) and severe (I_S_) symptomatic and hospitalized (H) with corresponding diagnosed states (D). All infections result in recovery (R) with partial immunity or fatality (F, D_F_); **B)** Contact matrix; **C)** Vaccination coverage under different rollout; **D)** Reactive social distancing in response to changes in hospitalization count; **E)** Variant replacement over time.

Major changes to previous model versions include separate compartments for mild and severe disease, age-specific relative susceptibility strata, vaccine efficacy against hospitalization, ability to handle multiple variants with increased transmissibility and virulence as well as differential vaccine efficacies against each variant. We now stratify the contact matrix by venue (school, home, work, other) and can adjust local mixing parameters to implement reactive reduction in contacts at each venue. We apply this to control transmission in schools by eliminating school contacts for the 2020-2021 school year, reflecting school closures, and considering a range of school reopening scenarios, from fully closed through open with reduced contacts to fully open for the 2021-2022 school year. Based on current state and county policies we assume that numbers of cases and hospitalizations are likely to fluctuate due to the community response to the epidemic[16]. When number of hospitalizations increase above 10 per 100,000 people per week, physical distancing measures are tightened, reducing contacts and thus transmission. Conversely, if the number of hospitalizations fall below 5 per 100,000 people per week physical distancing measures are gradually relaxed facilitating more contacts between susceptible and infected people. See Supplement for a complete description of the model and model equations.

We calibrated to data on diagnosed cases, hospitalizations, and deaths by age from King County using Approximate Bayesian Computation [17]. The calibration procedure was performed in two stages, first for the initial epidemic period in spring 2020 to calibrate the base transmission rate, reduction in transmission due to being diagnosed, and proportion of mild and severe cases, and second for the spring of 2021 to calibrate diagnostic rates for mild and severe illness, the relative proportion of asymptomatic cases diagnosed, and the current level of social distancing by age group. With this procedure, 100 parameterizations which minimized the distance between the normalized model outputs and normalized data for the 12 metrics (diagnosed cases, hospitalizations, and deaths for four age groups) were selected and used in the analysis (**Fig S2, Fig S3** and **Fig S4**). See Supplement for a complete description of the calibration procedure.

All simulations included in this analysis started on June 1^st^, 2021 with initial conditions calculated based on estimates of cumulative incidence, variant prevalence and vaccination coverage by age group as reported in King County. Cumulative incidence by age and current prevalence on June 1^st^, 2021 are back-calculated from hospitalization in King County through July 31^st^ while vaccination coverage by age group is derived from the CDC. The total prevalence by variant is set to the back calculated value for overall prevalence. Based on calibrated sets, approximately 80% of the infections on June 1, 2021 were with the Alpha variant while the remaining 20% are with Delta variant. Using this information, we populate all compartments (see Supplement for details).

### Simulated Scenarios

Our analysis is focused on the impact of vaccination programs based on: i) extending vaccination to children aged 5-11 with different starting dates (October 2021 and January 2022), ii) different proportions of physical interactions (PPI) at schools restored varying from completely closed to fully opened, and iii) improving the overall vaccination coverage among eligible population. The set of scenarios included in the main and additional analyses are listed in Table 2.

**Table 1.** Key model parameters.

| *Parameter description* | *Value* | *Source* |
| --- | --- | --- |
| **Maximum level of social distancing** by age (younger/older than 70) | (0.3/ 0.5) | [11] |
| **Minimum level of social distancing** by age (younger/older than 70) | (0.1/ 0.2) | [27] |
| **Threshold for triggering additional restrictions** on social interactions | more than 10 new hospitalizations per 100,000 individuals per week | [16,28] |
| **Threshold for releasing restrictions** on social interactions | fewer than 5 new hospitalizations per 100,000 individuals per week | [16,28] |
| **Vaccine efficacy on susceptibility** defined as reduction of the probability to acquire infection upon exposure by variant (Alpha, Delta) | (0.91, 0.82) | [5] [29] |
| **Vaccine efficacy on symptomatology** defined as reduction in the probability to develop symptoms upon infection by variant (Alpha, Delta) | (0.34, 0.34) | [5] |
| **Vaccine efficacy on hospitalization** defined as reduction in the risk of hospitalization for symptomatic infections by variant (Alpha, Delta) | (0.67, 0.67) | [30] |
| **Vaccine efficacy on infectiousness** defined as reduction in the probability to transmit different variants (Alpha, Delta) upon infection | (0,0) | Assumed |
| **Relative transmissibility** of variants (Alpha, Delta) compared to the original variant | (1.5, 2.4) | [31–34] |
| **Relative severity** of variants (Alpha, Delta) compared to the original variant | (1.5, 1.5) | [6,35,36] |
| **Proportion of infections which become symptomatic** by age group (0-19, 20-49, 50-69, 70+) | (0.25, 0.33, 0.55, 0.70) | [37] |
| **Proportion of symptomatic infections which are mild** and require no hospitalizations by age group (0-19, 20-49, 50-69, 70+) | (0.988-0.996, 0.96-0.995, 0.87-0.96, 0.62-0.87) | Calibrated |
| **Relative susceptibility** by age group (younger/older than 20) | (0.5/ 1) | [37] |

**Table 2.** Model parameters defining scenarios included in the main and sensitivity analyses (bold values are used in the main analysis)

| Parameter/Description | Values |
| --- | --- |
| **Proportions of physical interactions (PPI)** in schools relative to pre-COVID time | **0% (schools closed), 50%, 75%, 100%** |
| **Start of vaccinating kids ages 5-11** | **Oct 1, 2021, Jan 1, 2022, never** |
| **Overall vaccination coverage** achieved among subpopulation eligible for vaccination | 80%, **85%**, 90% |
| **Social distancing (SD) level**, defined as percentage reduction of COVID transmission due to limiting physical interactions including state and local mandates, masking, remote work, etc. | • **Reactive – varied within ranges listed in Table 1 in response to recorded number of hospitalizations over time**<br>• Fixed at minimum (10%) |
| **Waning of protection** against infection due to vaccination or prior infection | • **No waning**<br>• Waning to 50% of initial efficacy on susceptibility over 1 year without effect on the efficacy on hospitalization |

### Outcomes of interest

We measure the impact of different vaccination programs by comparing the cumulative hospitalizations (overall, by vaccination status and by age group), peak numbers of hospitalizations, the proportion of simulations requiring additional mitigation measures and the proportion of time under additional restrictions (SD max) calculated over the school year (Sept 2021 – June 2022). To study the impact of early vaccination of children aged 5-11 we also estimated the percentage reduction of hospitalizations over the same period compared to scenarios in which only individuals ages 12 and over are vaccinated.

## Results

Assuming that 85% of the currently eligible population (12 years and older) is vaccinated, implies that 72.9% of the total population is vaccinated with approximately 33% of the unvaccinated population being children of school age. If such vaccination coverage is achieved, our analysis shows that reopening schools in Fall 2021 will have a significant impact on expected SARS-COV-2 transmission in King County resulting in more hospital admissions over the school year. Our model projects 4945 (median, IQR 4622-5341) total COVID-19 hospitalizations if the physical contacts in schools are restored to their pre-COVID level (100% PPI) for the entire school year with 445 (median, IQR 418-473) hospitalizations at peak compared to 3675 (median, IQR 2311-4725) total and 324 (median, IQR 141-399) peak hospitalizations if schools remained closed **(Fig 2A and Fig 2B purple)**. Unregulated school reopening will likely result in 2-fold increases in pediatric hospitalizations from 163 to 325 (median, **Fig 3A, purple**) and require additional time under maximum social distancing imposed for 26% (median, IQR 23-28) of the school year to manage COVID outbreaks (**Fig 4A, purple**) with only 2% simulations avoiding restrictions compared to 62% for scenarios with schools closed (**Fig 4B, purple**). The majority of hospitalizations are projected to occur among unvaccinated people (see **Fig S5**) at per capita rate nearly 8-fold higher compared to vaccinated (median, **Fig 2C**).

**Figure 2.**
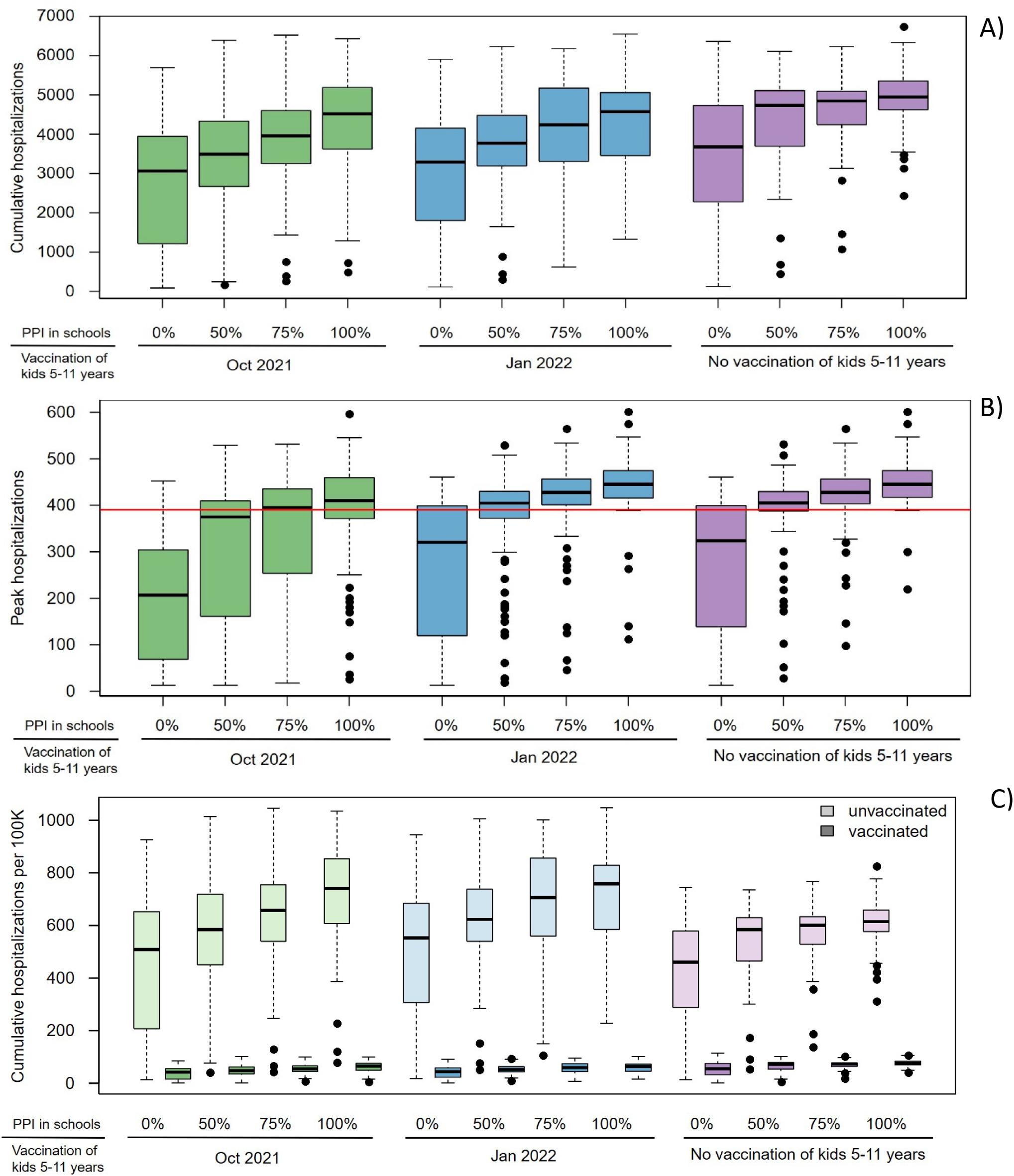
Overall hospitalizations expected under different scenarios of school reopening and extended vaccine eligibility: **A)** Cumulative hospitalizations over the 2021-2022 school year; **B)** The maximum number of people hospitalized with COVID at any given time over the school year and **C)** Overall per capita cumulative hospitalizations by vaccination status. Boxes represent interquartile range while whiskers extend to the most extreme data point which is no more than 1.5 times the interquartile range. Red line indicates 10% of the hospital bed capacity in King County used as a metric for public-health decisions.

**Figure 3.**
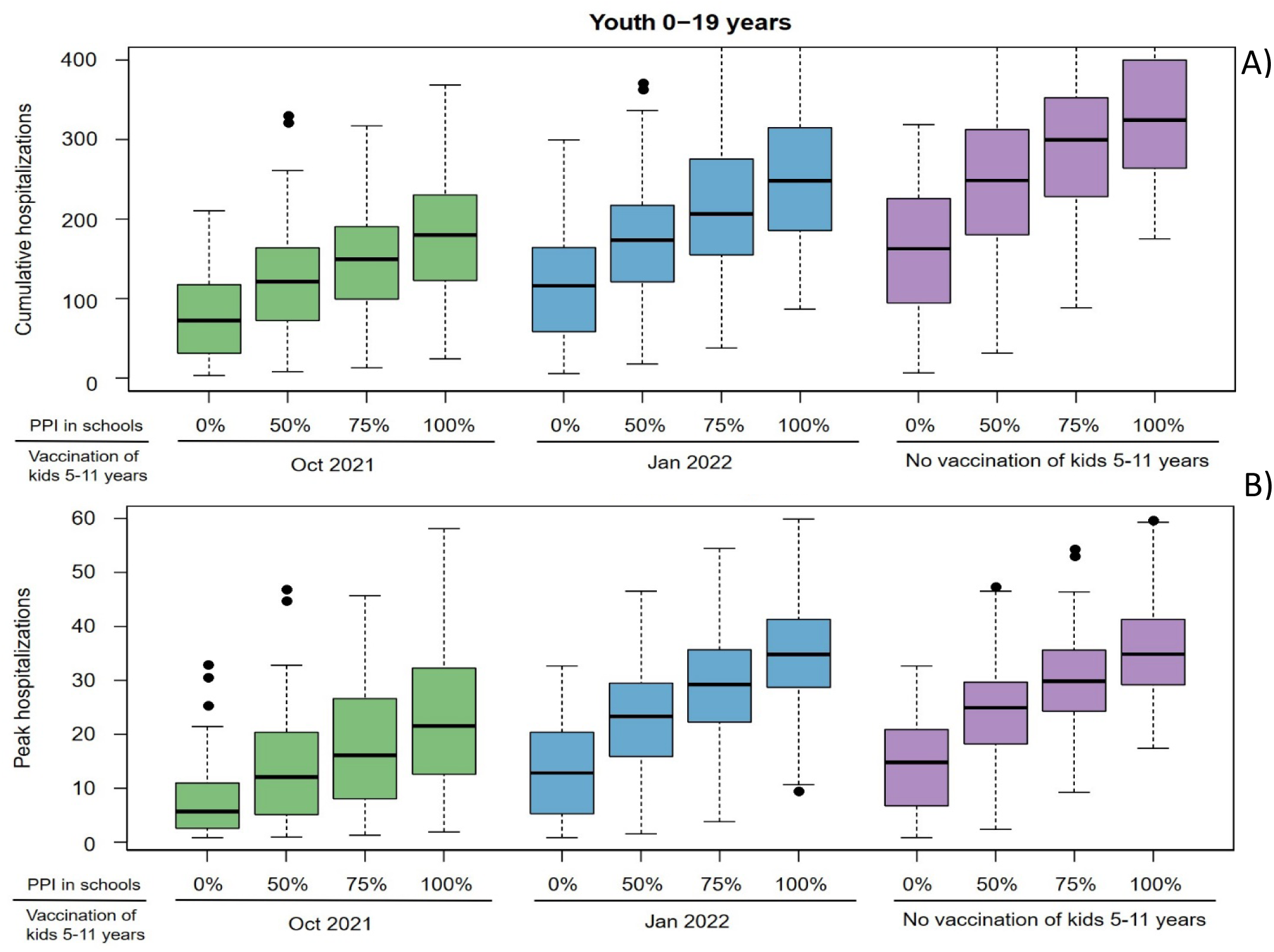
Hospitalizations among the youngest group (0-19 years) expected under different scenarios of school reopening and extended vaccine eligibility: **A)** Cumulative hospitalizations over the 2021-2022 school year and **B)** The maximum number of young people hospitalized with COVID at any given time over the school year.

**Figure 4.**
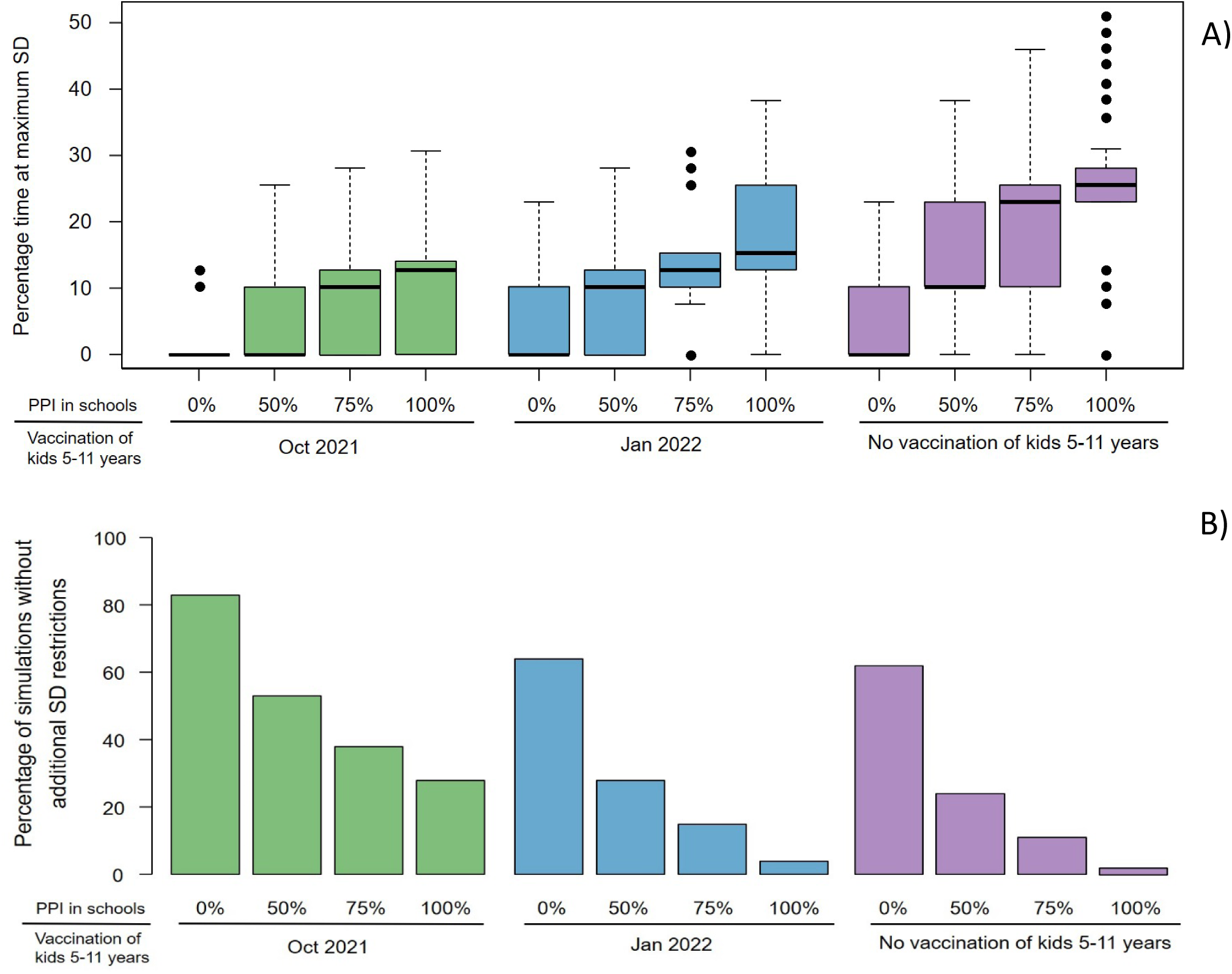
Effects of school reopening and extended vaccine eligibility on the need of additional COVID restrictions. A) Percentage time of the school year under maximum restricted social distancing (SD) and B) Percentage of simulations in which additional restrictions of social distancing are not required.

If kids age 5-11 are not vaccinated, reducing contacts at schools by 25% or 50% (through masking, ventilation and distancing) are expected to only slightly decrease the overall median cumulative hospitalizations (by 2% and 4% respectively, **Fig 2**) with more substantial effect on youth’s hospitalizations (median decreased by 8% and 23% respectively, **Fig 3**). Keeping 75% PPI in schools of the pre-COVID level will only shorten societal restrictions needed to keep COVID hospitalization rates below state-mandated thresholds by one week and increase the proportion of simulations that avoid restrictions to 11%. However, if physical contacts in schools are limited to 50%, the duration of societal restriction is likely to be cut in half and the proportion of simulations avoiding restriction would further double to ∼24% **(Fig 4)**.

Extending the vaccination eligibility to children aged 5-11 and achieving the same vaccination coverage (85%) in this age group would require approximately 150,000 additional vaccinations. It would increase the overall population vaccination coverage to 80.2% and significantly reduce the proportion of unvaccinated who are school age to 11%. As a result, between 400 and 1200 fewer hospitalizations are expected over the school year with vaccination of kids 5-11 starting in October 2021 across scenarios with different levels of PPI in schools **(Fig 2A green)**. Regardless of the level of physical interaction at school, the expected number of hospitalizations in the youngest age group is cut roughly half. Notably, significantly more youth hospitalizations are prevented if physical contacts in schools are fully restored (100% PPI) or in-person schooling is managed with some mitigation measures (75% PPI) (**Fig S6B**, green). Moreover, offering early vaccination to children aged 5-11 will reduce the need of additional social distancing **(Fig 4 green)**. For instance, in the scenario with 75% PPI in schools, the median number of hospitalizations over the school year is projected to be 13% lower than if school contacts are fully restored (100% PPI) with only slight reduction in the expected time under societal restrictions. However, controlling physical contacts in schools to 50% of the pre-COVID level (50% PPI) will further reduce the expected median number of hospitalizations by 23% and increase the likelihood of avoiding societal restrictions above 50%. It will also likely keep the proportion of hospital beds filled with COVID patients below the county target of 10% **(Fig 2B green)** [16]. Delaying childhood vaccination by 3 months will void most of the benefits measured over the school year including reductions in total, pediatric hospitalizations and days requiring maximal social distancing. **(Fig 2, 3 and 4, blue)**

Next, we explore the effects of improved vaccination coverage in King County. Increasing the proportion of vaccinated individuals aged 12+ to 90% will require approximately 100,000 additional vaccinations and is projected to have a significant effect on the expected hospitalizations over the school year across all scenarios. If children aged 5-11 remain unvaccinated with 75% PPI in schools, reaching 90% overall vaccination coverage will prevent on average approximately 1300 hospitalizations or one quarter of the projected **(Fig 5A purple)** but still almost 50% of the simulations require additional restrictions **(Fig 5B purple)**. The benefit of vaccinating 90% of the eligible population is largest when combined with early vaccination of children aged 5-11. In those scenarios it is projected to prevent ∼2400 hospitalizations on average over the school year which is 60% of those estimated with 85% coverage **(Fig 5A green)** and almost certainly would avoid the need of extra periods of social distancing **(Fig 5B green)**. Conversely, if the overall vaccination coverage remains at the level reached in Sept 2021 (80%) we project more hospitalizations across scenarios and very slim chance to avoid additional societal restrictions. For instance, with school contacts at 75% of pre-pandemic levels, our model projects approximately 600-1200 more hospitalizations compared to simulations with 85% coverage **(Fig 5 blue)**.

**Figure 5.**
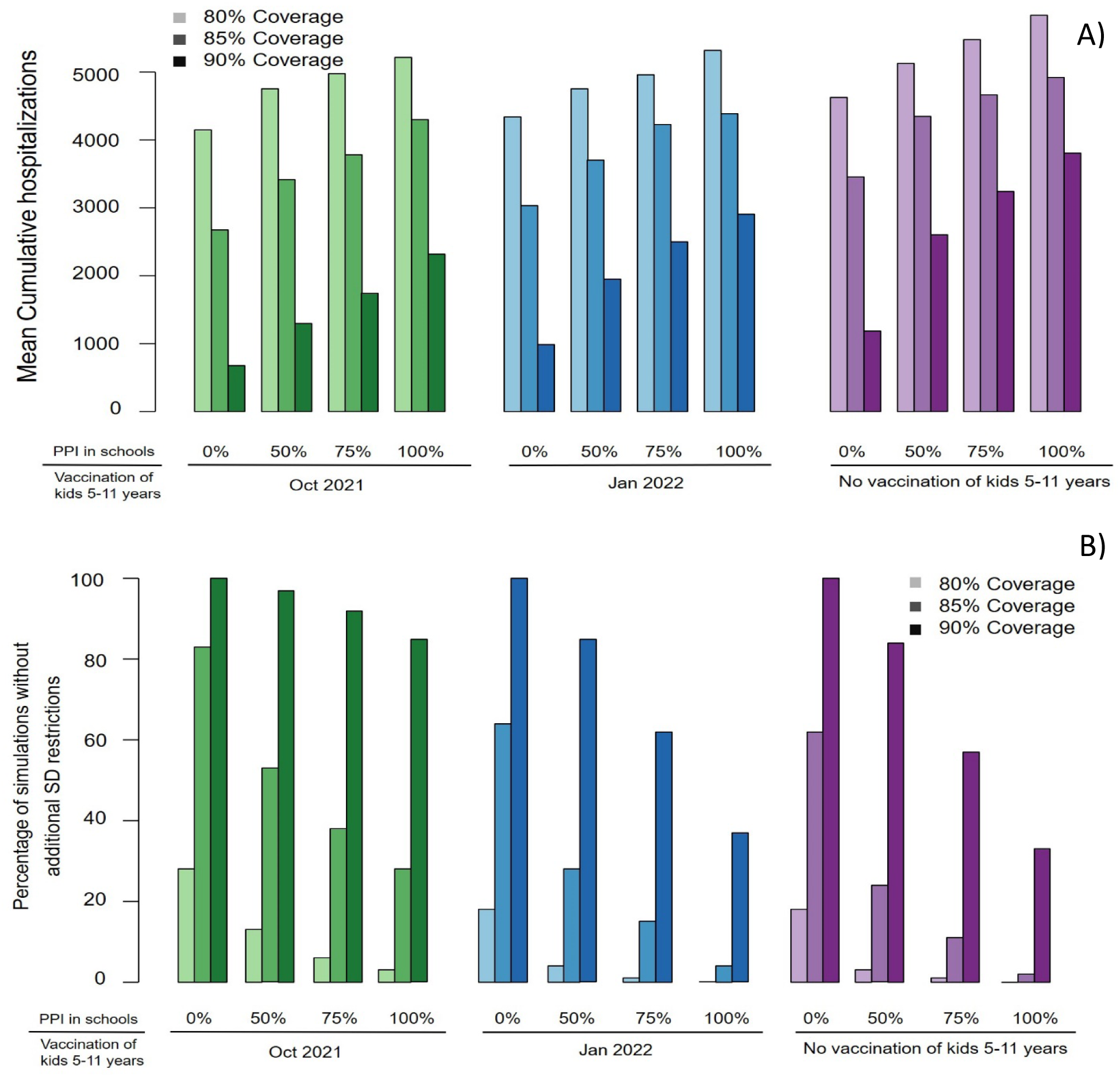
Importance of achieving better vaccination coverage. **A)** Mean **c**umulative hospitalizations over the school year and **B)** Percentage of simulations in which additional restrictions of social distancing are not required.

### Simulations without reactive social distancing

We explored the influence of our assumptions on reactive social distancing by simulating alternative scenarios in which the social interactions were kept at the lowest level throughout the school year with no dynamic adjustments in response to increasing number of hospitalizations **(Fig 6)**. The results suggest that reactive social distancing is most important for scenarios with 100% PPI in schools and no vaccination for children aged 5-11 where the number of hospitalizations is expected to increase by 123% on average. If all school-aged kids are eligible for early vaccination the projected increase in absence of reactive distancing is limited to 65% **(Fig 6 green)** while if schools remain closed for the school year this increase is limited to 28%.

**Figure 6.**
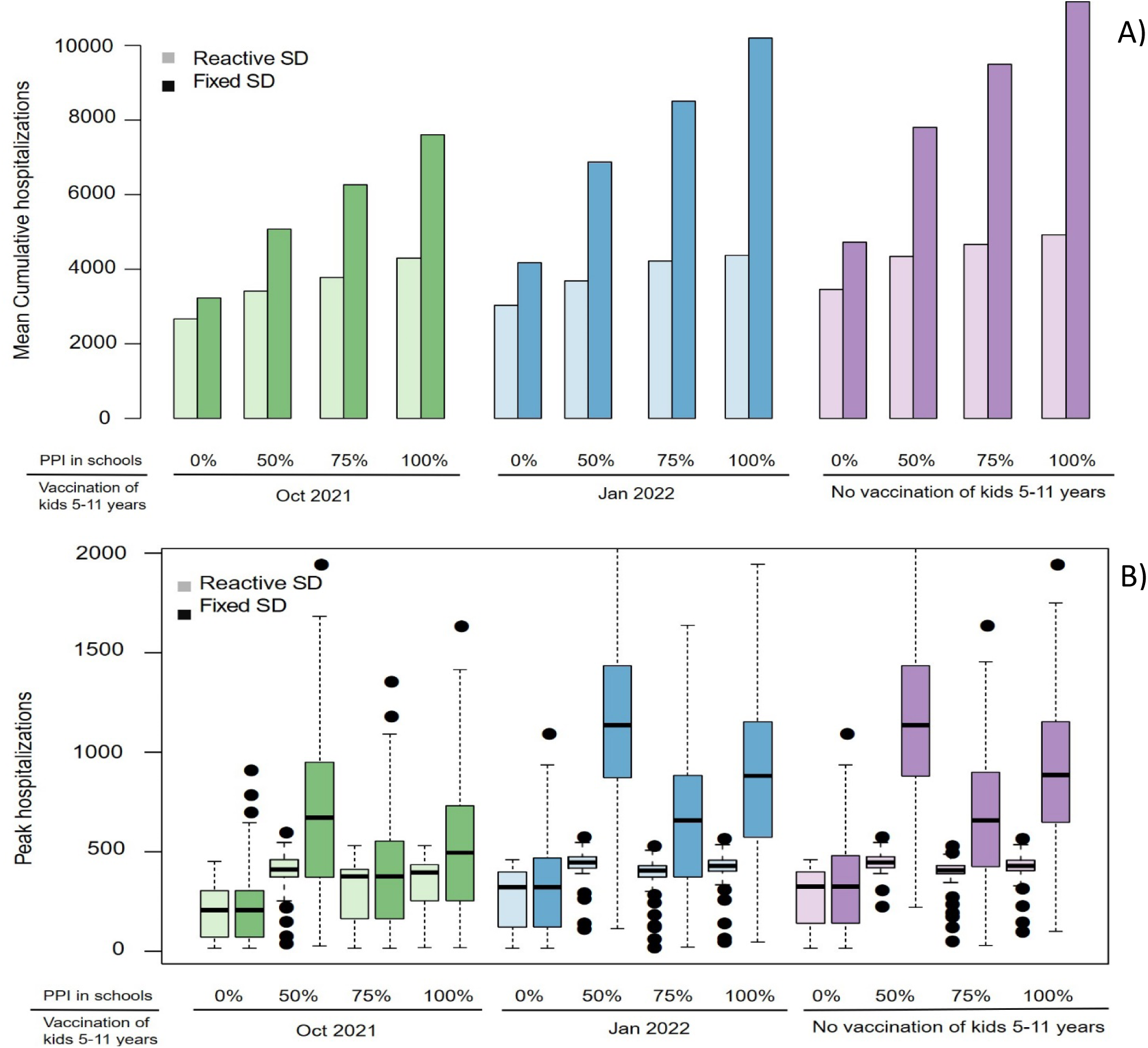
Importance of reactive social distancing. **A)** Mean cumulative hospitalizations and **B)** peak hospitalization over the school year under scenarios with reactive social distancing compared to scenarios in which restrictions are kept at the minimum level.

### Simulations with immune waning

We also simulated scenarios if protection against infection provided by vaccine or previous infection decreases by half over an average of a year while preserving protection against hospitalization **(Fig S7)**. Our model projected: i) between 8000 and 10,000 hospitalizations under these scenarios which is a 2- to 3-fold increase over projections from our main analysis; ii) large increase of the expected peak number of hospitalizations up to 650-750 and iii) additional social restrictions needed for more than 75% of the school year. These results suggest that more stringent restrictions of social interactions combined with mass vaccine boosting may be necessary.

## Discussion

Our simulations suggest that due the highly infectious nature of the SARS-CoV-2 Delta variant, keeping schools open with contacts reestablished at pre-pandemic levels (no in-school masking, physical distancing or testing) would result in unmanageable numbers of hospitalizations, even in communities with high vaccination rates. Therefore, continual maintenance of non-pharmaceutical interventions among younger cohorts will be necessary to limit the scope of further infections as vaccine rollout continues slowly. These results agree with previous work and have been largely validated by the current experiences across the US. [12,18,19] In particular, states with low vaccination rates where all non-pharmaceutical interventions have been lifted have seen the largest number of hospitalizations in recent weeks that have put their healthcare systems in at the brink of collapse with low ICU and overall bed availability. [20–22] Further, our results align with other recent modeling results suggesting that without any school-based interventions, the majority of school-age students would become infected during the school year [12]. Lessler et al showed that seven simultaneous mitigation strategies were needed to be put in place at schools to significantly reduce school-associated risk. [23] Consistent with this finding, we suggest that reducing potentially infectious contacts to 50% of pre-pandemic levels would result in a significant reduction of hospitalizations and infections in school-age children.

Pediatric cases in the US increased exponentially in the first weeks of September 2021, and hospitalizations for children and adolescents increased 5-fold between June and mid-August with the rise of the Delta variant, prompting the American Academy of Pediatrics to plead with the FDA to accelerate vaccine approval for the pediatric population. [24,25] Pfizer recently announced favorable preliminary results for the clinical trials for their vaccine in children aged 5 to 11. [8] We demonstrate that vaccination of this age group as soon as possible would further reduce the burden of COVID-19 disease in children, averting up to 60% hospitalizations in that age group. Further, our simulations suggest that delaying vaccination start to January 1^st^ 2022 would lead to a far less substantial impact on the current Delta wave. Children are the only age group for whom the vaccine is not approved, and currently they are reliant on others to be protected. Our simulations suggest that in-person schooling will result in large numbers of infections in this age group. It is an urgent priority to provide children with a safe and effective vaccine to allow them to continue their education while being directly protected.

Finally, we demonstrate the continued importance of targeting the remaining unvaccinated population of adults for vaccination. Even 5% increases in vaccination coverage among this group would result in a disproportionate reduction in hospitalizations and could eliminate the need for any periods of required further social distancing. Although seemingly small, such improvement may require promoting all best practices at the state level to be achieved, given that the vaccination coverage in King County is already high. However, data from countries like Ireland with 90.3% of the adult population vaccinated including close to 100% of seniors (65+) is encouraging. [26]

Each of these conclusions become even more urgent when we consider the possibility of immune waning for previously infected and vaccinated people. Under this assumption, hospitalizations rise among all age groups and it is likely that boosting will be required to limit the spread of virus in the community.

As with any simulation-based analysis, our work has several limitations. We assume vaccine protects vaccinated individuals immediately after vaccination, and we do not explicitly model two-dose vaccination. Our model is stratified by age, and we do not consider other factors such as occupation, socio-economic status and race that are known to affect both susceptibility to infection and to severe disease. Our model is not a forecast tool for the specific reason that the multifactorial components of social distancing cannot be measured or predicted. Our model is not intended to make specific projections for other regions or counties where vaccination rates, masking, age structure, and SARS-CoV-2 seroprevalence differ. However, the model’s qualitative conclusions are likely to be highly relevant for most communities.

Overall, our work highlights three main messages. First, nonpharmaceutical interventions must be maintained vigilantly in schools to prevent both pediatric and adult hospitalizations. Second, vaccination of school-aged children will only exert a meaningful impact on the epidemic over this school year if implemented rapidly within the next one to two months. Finally, it remains critical to vaccinate as many unvaccinated adults as possible to limit the morbidity and mortality associated with the current surge in Delta variant cases.

## Supporting information

Technical Supplement

## Data Availability

All modeling results are available upon request.

## Acknowledgements

This work was partially supported by grants from the National Institute of Allergy and Infectious Diseases of the National Institutes of Health (UM1AI068635) and (R01 AI121129-04 to Dr. Schiffer). CB, MM, DAS, JTS and DD were also supported by a grant from Centers for Disease Control and Prevention (NU38OT000297-02) through their cooperative agreement with the Council of State and Territorial Epidemiologists. DBR reported support from the Washington Research Foundation. Scientific Computing Infrastructure at Fred Hutch was funded by ORIP grant S10OD028685.

## References

1. Centers for Disease Control and Prevention. Interim Public Health Recommendations for Fully Vaccinated People. Available at: https://www.cdc.gov/coronavirus/2019-ncov/vaccines/fully-vaccinated-guidance.html#anchor_1619526549276. Accessed 21 September 2021.

2. Centers for Disease Control and Prevention. Covid Data Tracker : Variant Proportions.

3. Washington State Department of Health. SARS-CoV-2 Sequencing and Variants in Washington State Washington State Department of Health. 2021. Available at: https://www.doh.wa.gov/Portals/1/Documents/1600/coronavirus/data-tables/420-316-SequencingAndVariantsReport.pdf.

4. Campbell F, Archer B, Laurenson-Schafer H, et al. Increased transmissibility and global spread of SARSCoV-2 variants of concern as at June 2021. Eurosurveillance 2021; 26:1–6.

5. Sheikh A, McMenamin J, Taylor B, Robertson C. SARS-CoV-2 Delta VOC in Scotland: demographics, risk of hospital admission, and vaccine effectiveness. Lancet 2021; 397:2461–2462.

6. Fisman D, Tuite A. Progressive Increase in Virulence of Novel SARS-CoV-2 Variants in Ontario, Canada. 2021.

7. Tartof SY, Slezak JM, Fischer H, et al. Six-Month Effectiveness of BNT162B2 mRNA COVID-19 Vaccine in a Large US Integrated Health System: A Retrospective Cohort Study. Available at: https://papers.ssrn.com/sol3/papers.cfm?abstract_id=3909743.

8. Pfizer BioNTech. Pfizer and BioNTech Announce Positive Topline Results From Pivotal Trial of COVID-19 Vaccine in Children 5 to 11 Years. Available at: https://www.businesswire.com/news/home/20210920005452/en/.

9. King County. Summary of COVID-19 vaccination among King County residents. Available at: https://kingcounty.gov/depts/health/covid-19/data/vaccination.aspx. Accessed 21 September 2021.

10. Washington Governor Jay Inslee. Inslee signs emergency proclamation requiring in-person education opportunities for public K-12 schools. Available at: https://www.governor.wa.gov/news-media/inslee-signs-emergency-proclamation-requiring-person-education-opportunities-public-k-12. Accessed 27 September 2021.

11. Bracis C, Burns E, Moore M, et al. Widespread testing, case isolation and contact tracing may allow safe school reopening with continued moderate physical distancing: A modeling analysis of King County, WA data. Infect Dis Model 2021; 6:24–35. Available at: https://doi.org/10.1016/j.idm.2020.11.003.

12. Zhang Y, Johnson K, Yu Z, et al. COVID-19 Projections for K12 Schools in Fall 2021: Significant Transmission without Interventions. medRxiv 2021; :2021.08.10.21261726. Available at: http://medrxiv.org/content/early/2021/09/03/2021.08.10.21261726.abstract.

13. Reeves DB, Bracis C, Swan DA, et al. Rapid vaccination and partial lockdown minimize 4th waves from emerging highly contagious SARS-CoV-2 variants. Med 2021; 2:573–574. Available at: https://doi.org/10.1016/j.medj.2021.04.012.

14. Swan DA, Bracis C, Janes H, et al. COVID-19 vaccines that reduce symptoms but do not block infection need higher coverage and faster rollout to achieve population impact. Sci Rep 2021; 11:1–9.

15. Swan DA, Goyal A, Bracis C, et al. Vaccines that prevent SARS-CoV-2 transmission may prevent or dampen a spring wave of COVID-19 cases and deaths in 2021. Viruses 2021;

16. King County. Key indicators of COVID-19 activity in King County. https://kingcounty.gov/depts/health/covid-19/data.aspx

17. Beaumont MA, Cornuet JM, Marin JM, Robert CP. Adaptive approximate Bayesian computation. Biometrika 2009; 96:983–990.

18. Jehn M, Mccullough J Mac, Dale AP, et al. Association Between K – 12 School Mask Policies and School-Associated COVID-19 Outbreaks — Maricopa and Pima Counties, Arizona, July – August 2021. Morb Mortal Wkly Report, US Dep Heal Hum Serv Dis Control Prev 2021; 70:2020–2022.

19. Budzyn SE, Panaggio MJ, Parks SE, Papazian M, Magid J, Barrios LC. Pediatric COVID-19 Cases in Counties With and Without School Mask Requirements — United States, July 1 – September 4, 2021. Morb Mortal Wkly Report, US Dep Heal Hum Serv Dis Control Prev 2021; 70:21–23.

20. These 5 states have less than 10% of ICU beds left as Covid-19 overwhelms hospitals. Available at: https://www.cnn.com/2021/08/31/health/us-coronavirus-tuesday/index.html.

21. Mississippi’s Hospital System Could Collapse Within 10 Days Under COVID’s Strain. Available at: https://www.npr.org/2021/08/12/1027103023/florida-mississippi-arkansas-hospitals-overwhelmed-covid-19-delta.

22. Health and Human Services. Hospital Utilization. Available at: https://protect-public.hhs.gov/pages/hospital-utilization.

23. Lessler J, Grabowski MK, Grantz KH, et al. Household COVID-19 risk and in-person schooling. Science (80-) 2021; 2939:1–10.

24. American Academy of Pediatrics, Children’s Hospital Association. Children and COVID-19: State Data Report Version: 6/24/21. 2021. Available at: https://downloads.aap.org/AAP/PDF/AAP and CHA - Children and COVID-19 State Data Report 6.24 FINAL.pdf.

25. Delahoy MJ, Ujamaa D, Whitaker M, et al. Hospitalizations Associated with COVID-19 Among Children and Adolescents - COVID-NET, 14 States, March 1, 2020–August 14, 2021. Morb Mortal Wkly Report, US Dep Heal Hum Serv Dis Control Prev 2021; 70:1255–1260.

26. Covid-19 Vaccine Tracker: How many people have been inoculated in Ireland?

27. Abaluck J, Kwong LH, Styczynski A, et al. The Impact of Community Masking on COVID-19 : A Cluster-Randomized Trial in Bangladesh. 2021. Available at: https://www.poverty-action.org/publication/impact-community-masking-covid-19-cluster-randomized-trial-bangladesh.

28. Washington State Department of Health. Healthy Washington : Roadmap to Recovery Report. 2021. Available at: https://www.doh.wa.gov/Portals/1/Documents/1600/coronavirus/data-tables/420-316-SequencingAndVariantsReport.pdf.

29. Bernal JL, Andrews N, Gower C, et al. Effectiveness of COVID-19 vaccines against the B.1.617.2 variant. 2021. Available at: https://www.medrxiv.org/content/10.1101/2021.05.22.21257658v1.full.pdf.

30. Stowe J, Andrews N, Gower C, et al. Effectiveness of COVID-19 vaccines against hospital admission with the Delta (B.1.617.2) variant. 2021.

31. Davies NG, Abbott S, Barnard RC, et al. Estimated transmissibility and impact of SARS-CoV-2 lineage B.1.1.7 in England. Science (80-) 2021; 372:eabg3055.

32. Volz E, Mishra S, Chand M, et al. Assessing transmissibility of SARS-CoV-2 lineage B.1.1.7 in England. Nature 2021; 593:266–269.

33. Dhar AMS, Marwal R, Vs R, Ponnusamy K. Genomic characterization and Epidemiology of an emerging SARS-CoV-2 variant in Delhi, India Affiliations : medRxiv 2021; :1–17.

34. Public Health England. SARS-CoV-2 variants of concern and variants under investigation in England. 2021.

35. Dagpunar J. Interim estimates of increased transmissibility, growth rate, and reproduction number of the Covid-19 B.1.617.2 variant of concern in the United Kingdom. medRxiv 2021;

36. Ong SWX, Chiew CJ, Ang LW, et al. Clinical and virological features of SARS-CoV-2 variants of concern: a retrospective cohort study comparing B.1.1.7 (Alpha), B.1.315 (Beta), and B.1.617.2 (Delta). Clin Infect Dis 2021; Available at: https://academic.oup.com/cid/advance-article/doi/10.1093/cid/ciab721/6356459.

37. Davies NG, Klepac P, Liu Y, et al. Age-dependent effects in the transmission and control of COVID-19 epidemics. Nat Med 2020; 26:1205–1211.

