## Supplementary material for "Improving vaccination coverage and offering vaccine to all school-age children will allow uninterrupted in-person schooling in King County, WA: Modeling analysis": Technical Supplement

### 1. Model description

To model SARS-CoV-2 epidemiological dynamics, we have developed an extended SIR model. The model is (1) age structured, (2) contains vaccinated, unvaccinated, and recovered susceptible individuals, and (3) allows for the introduction of new viral variants (also referred to as strains). Therefore, each state variable can now be expressed as a tensor $\mathbf{X} = X(a, v, q)$ for state X and age $a$, vaccination status $v$, and strain $q$. Some rates also depend on these variables. Importantly, these variables can each have a flexible number of possible values. That is, the default version of the model has four age classes, but can defined with any given $n$ age groups, including just one age group for a non-age-structured model. Similarly, for vaccination, $v = v_0$ corresponds to an unvaccinated state and the model supports an arbitrary $p$ number of vaccines, and likewise for variants an arbitrary $m$ number can be defined. Finally, an arbitrary number of recovered values can be defined, which functions as 'quasi-vaccinated' values which also provide partial protection against reinfection. Because the length of $\mathbf{S}$ is now different than other states, the matrix M converts between $\mathbf{S}$ and other states by summing across strains.

The number of individuals (across age $a$, vaccination status $v$, and strain $q$) in each compartment can then be calculated as sums, e.g. $\sum_{a,v,q} X(a, v, q)$. By summing only over certain variables, we can access quantities such as the number of vaccinated infected adults with B.1.1.7, or the number of recovered children who were infected with the original 2020 circulating strain.

The model also has the flexibility to make any parameter time-varying. After setting the initial values, there is a framework for specifying the date and the new value for the parameter. This is used, for example, with mortality rates, which were higher early in the epidemic and then declined as treatments improved. Similarly, it is possible to calibrate a set of values for a parameter with specified time periods. This capability is used foremost for social distancing, which varies consider-able throughout the epidemic, but also for parameters like diagnosis rate which are known to differ throughout epidemic.

In addition, we have a publicly available tool https://covidmodeling.fredhutch.org/ which has additional documentation and the ability for the user to see the most up to date projections and explore many variables in the 'user-adjustable' page.

**2. Model structure** The model is expressed as the set of differential equations (where overdot denotes time derivative), with states listed in Supplementary Table 1 and parameters in Supplementary Table 2,

$$
\begin{aligned}
\dot{\mathbf{S}} &= -M^T \times \lambda(a,v,q)\mathbf{S} - M^T \times \nu(a,q) \\
&\quad + Y \times (r_M\mathbf{A2} + r_M\mathbf{IM} + r_H\mathbf{H} + r_A\mathbf{DA2} + r_M\mathbf{DM} + r_H\mathbf{DH}) \\
&\quad + W \times \mathbf{S} \\
\dot{\mathbf{E}} &= \lambda(a,v,q)M \times \mathbf{S} - \gamma_1\mathbf{E} + \nu(a,q) \\
\dot{\mathbf{A1}} &= (1 - \{1 - \mathrm{VE}_{\mathrm{SYMP}}(v,q)\}\,\pi)\gamma_1\mathbf{E} - (\rho_A\Delta_{IM}(a) + \gamma_2)\mathbf{A1} \\
\dot{\mathbf{A2}} &= \gamma_2\mathbf{A1} - (\rho_A\Delta_{IM}(a) - r_A)\mathbf{A2} \\
\dot{\mathbf{P}} &= \pi\,\{1 - \mathrm{VE}_{\mathrm{SYMP}}(v,q)\}\,\gamma_1\mathbf{E} - (\rho_A\Delta(a) + \gamma_2)\mathbf{P} \\
\dot{\mathbf{IM}} &= \gamma_2 m(a)\mathbf{P} - (\Delta_{IM}(a) + r_M)\mathbf{IM} \\
\dot{\mathbf{IS}} &= \gamma_2(1 - m(a))\mathbf{P} - (\Delta_{IS}(a) + h(a) + f(a))\mathbf{IS} \\
\dot{\mathbf{H}} &= h(a)\mathbf{IS} - (r_H + \delta_H + f(a))\mathbf{H} \\
\dot{\mathbf{F}} &= f(a)(\mathbf{H} + \mathbf{IS}) \\
\dot{\mathbf{DA1}} &= \rho_A\Delta_{IM}(a)\mathbf{A1} - \gamma_2\mathbf{DA1} \\
\dot{\mathbf{DA2}} &= \gamma_2\mathbf{DA1} + \rho_A\Delta_{IM}(a)\mathbf{A2} - r_A\mathbf{DA2} \\
\dot{\mathbf{DP}} &= \rho_A\Delta(a)\mathbf{P} - \gamma_2)\mathbf{DP} \\
\dot{\mathbf{DM}} &= \gamma_2 m(a)\mathbf{DP} + \Delta_{IM}(a)\mathbf{IM} - r_M\mathbf{DM} \\
\dot{\mathbf{DS}} &= \gamma_2(1 - m(a))\mathbf{DP} + \Delta_{IS}(a)\mathbf{IS} - (h(a) + f(a))\mathbf{DS} \\
\dot{\mathbf{DH}} &= h(a)\mathbf{DS} + \delta_H\mathbf{H} - (r_H + f(a))\mathbf{DH} \\
\dot{\mathbf{DF}} &= f(a)(\mathbf{DH} + \mathbf{DS})
\end{aligned}
\tag{1}
$$

A schematic cartoon of the model is provided as Supplementary Figure 1. In words, this model describes a population of susceptible individuals $S(a, v = v_0, q)$, who can become vaccinated $S(a, v \in v_1, ..., v_p, q)$ and/or infected. Infected individuals begin in an exposed state $\mathbf{E}$, and after a waiting time $\gamma_1$ proceed to infection as asymptomatic ($\mathbf{A1}$, probability $1 - \pi$) or symptomatic ($\mathbf{P}$, probability $\pi$). Note that $\mathbf{P}$ is a pre-symptomatic period with waiting time $\gamma_2$ for symptomatic infections. After the pre-symptomatic period, symptomatic infections are divided into mild ($\mathbf{IM}$, probability $m(a)$) or severe ($\mathbf{IS}$, probability $1 - m(a)$) infections. Mild infections recover with rate $r_M$. Asymptomatic infections have an analogous state $\mathbf{A2}$ from which they recover at rate $r_A$. Severe infections are defined as those that lead to hospitalization, $\mathbf{H}$, and possibly death, $\mathbf{F}$. Hospitalized infected recover at rate $r_H$, or die with rate $f(a)$ and it is also possible to die outside the hospital with the same rate.

Infected individuals are diagnosed as COVID-19 cases with a rate $\Delta_X(a)$ that varies by symptom severity ($\Delta_{IM}(a), \Delta_{IS}(a), \Delta_H$ for mild, severe, and hospitalized respectively. Asymptomatic infections are assumed to be diagnosed at a rate proportionally inferior to symptomatic defined by $\rho_A$. Diagnosed individuals move to a set of parallel states, $\mathbf{DA1}$, $\mathbf{DA2}$, $\mathbf{DP}$, $\mathbf{DM}$, $\mathbf{DS}$, $\mathbf{DH}$ and $\mathbf{DF}$. We assume any individual who dies was diagnosed ultimately. That is, the states $\mathbf{F}$ and $\mathbf{DF}$ are combined for the total number of deaths. Diagnosed cases are used for comparing to data and for lowering transmission among diagnosed (see below).

Recovery is not a separate state, but incorporated into the susceptible state to allow for reinfection (immunity can be parameterized to be partial or complete). The number of recovered states is also flexible. For example, information on infecting strain can be retained to investigate cross-strain immunity, or vaccination status can be assumed to trump infection in terms of re-infection, or other possibilities. How individuals move from the infected states to the compartments of **S** are described by the Y matrix. Waning immunity, that is moving individuals from the vaccinated and recovered compartments of **S** to be newly sussceptible, is also possible and is described by the W matrix.

Vaccinations occur by adjusting the status of susceptible (or recovered, because recovered individuals can still be vaccinated) within **S**. Infected individuals with new variants can enter the system with rate $\nu_{import}(a, q)$. Note we assume a closed system (conservation of number), meaning this model could be interpreted as some KC resident visited elsewhere, contracted the variant, and returned or also that some KC resident left permanently precisely when a newly infected individual entered. New strains are also defined by their infectivity $\nu_{inf}(q)$ and severity $\nu_{sev}(a, q)$, relative to the original strain. It is also possible to set the starting prevalence of different strains in the initial conditions, which we favor as this is generally more straightforward to estimate than the importation rate.

There are several additional model features that adjust the dynamical system, which are detailed in the following sections.

**Supplementary Table 1.** Table of model states.

| State | Description | Notes |
|---|---|---|
| S | Susceptible | Different length than other states because no infecting strain |
| E | Exposed | |
| A1 | Asymptomatic 1 | Analog to P |
| A2 | Asymptomatic 2 | Analog to IM/IS |
| P | Pre-symptomatic | |
| IM | Infected (mild) | |
| IS | Infected (serious) | |
| H | Hospitalized | |
| F | Dead | Assumed diagnosed after death |
| DA1 | Diagnosed Asymptomatic 1 | Analog to DP |
| DA2 | Diagnosed Asymptomatic 2 | Analog to DM/DS |
| DP | Diagnosed Pre-symptomatic | |
| DM | Diagnosed Infected (mild) | |
| DS | Diagnosed Infected (serious) | |
| DH | Diagnosed Hospitalized | |
| DF | Diagnosed Dead | |

***Force of infection.***

The force of infection $\lambda(a, v, q)$ governing transmission depends on the state of the transmitting individual (e.g. asymptomatic transmission is less-likely, certain variants are more infectious, certain ages interact less). It also depends on vaccination and the time-dependent reduction in contacts mediated by social/physical distancing $\sigma(a)$.

$$\begin{aligned}\lambda(a, v, q) =&[1-\sigma(a)](1-\mathrm{VE}_{\mathrm{SUSC}}(v, q))\kappa(a)\\ &\mathcal{C}(a) \times (1-\mathrm{VE}_{\mathrm{INF}}(v, q))\beta^{*}[1-\sigma(a)] \sum_{X \in \mathcal{X}} \beta_X X(a, v, q)/N(a)\end{aligned} \tag{2}$$

where $\mathcal{X} = \{A1, A2, P, IM, IS, H, DA1, DA2, DP, DM, DS, DH\}$ is the set of all potentially infectious states. Naturally, susceptible, exposed, recovered, and deceased individuals also do not contribute to ongoing infection.The key parameter for infection is $\beta*$, the base rate of transmission, which is further modified by state-specific transmission rates, $\beta_A 1$, $\beta_A 2$, $\beta_P$, $\beta_M$, $\beta_S$, $\beta_H$. Note it is assumed that hospitalized individuals do not contribute to transmission ($\beta_H = 0$). For the diagnosed states, the effect of reducing interactions upon being diagnosed is handled by multiplying the state-specific $\beta_X$ by the reduction in transmission due to diagnosis, $\beta_D$. Both $\beta*$ and $\beta_D$ are calibrated parameters.

The model uses an empirically derived contact matrix $\mathcal{C}(a)$ that parameterizes the probability of interactions between transmitting and exposed individuals in different age groups. The contact matrix is assembled from those specifying interactions from different locations, and a location-specific social distancing parameter can also be specified. Note that these location-specific social distancing parameters are scalars and not age-stratified.

$$\begin{aligned}\mathcal{C}(a) =&(1-\sigma_{school})\mathcal{C}_{school}(a) + (1-\sigma_{home})\mathcal{C}_{home}(a)\\ &+ (1-\sigma_{work})\mathcal{C}_{work}(a) + (1-\sigma_{other})\mathcal{C}_{other}(a)\end{aligned} \tag{3}$$

Transmission is further affected by $\kappa(a)$ the relative age-specific susceptibility. The reduction in transmission due to social distancing is handled by the $\sigma(a)$, which varies between 0 and 1 and is also age-specific (i.e., the more susceptible oldest age group can have a higher value for social distancing than younger age groups). It is assumed that social distancing affects both transmission and susceptibility, thus it is applied to both sides of the contact matrix $\mathcal{C}(a)$. The location-specific social distancing parameters ($\sigma_{school}$, $\sigma_{home}$, $\sigma_{work}$, and $\sigma_{other}$) reduce only interactions corresponding to that location in the contact matrix, for example due to the closing of schools.

*Reactive social distancing*. We include a time-varying, age-stratified vector $\sigma(a, t)$ that governs social distancing (non-pharmaceutical interventions) including reduced contacts through personal choices and/or mandated partial lockdowns, as well as reductions in exposure contacts due to mask wearing, physical distancing, capacity limitations, vaccination requirements, etc. $\sigma(a, t)$ varies from 0, indicating pre-pandemic levels of societal interactivity and no masking, to 1, indicating complete lockdown with no interactions. $\sigma(a, t)$, along with reductions in the susceptible population due to infection or vaccination, is the main driver of $R_{effective}$ and therefore epidemic peaks and declines.

After the calibration period, the issue is how to set $\sigma(a,t)$ for forward simulation into the future, as $\sigma(a,t)$ is changing through time in response to human behavior. We use dynamical social distancing based on a specified threshold, such as the current diagnosed cases or hospitalizations or some combination. The threshold is a flexible, user-specified function, and thus can be based on government criteria for implementing restrictions, such as weekly or bi-weekly case or hospitialization numbers. This function is parameterized by the following values: the maximum $T_{\max}$ and minimum $T_{\min}$ thresholds for increasing or decreasing restrictions (corresponding to changing $\sigma$), the period $\tau$ over which the threshold is calculated, the restricted and released social distancing values $\sigma_{\max}(a)$ and $\sigma_{\min}$, and the increment $\sigma_{inc}$ by which to change $\sigma$ when releasing restrictions. Thus we have

$$\sigma(a,t) = \begin{cases} \sigma_{\max}(a) & \sum_\tau \mathbf{T} > T_{\max} \\ \sigma(a,t) - \sigma_{inc} & \sum_\tau \mathbf{T} < T_{\min} \\ \sigma_{\min} & \sigma(a,t) - \sigma_{inc} < \sigma_{\min} \end{cases} \tag{4}$$

where the sum of the threshold metric(s) $\sum_\tau \mathbf{T}$ is taken over time period $\tau$. Based on local policy for decision making, this is set at $\tau = 1$ week intervals in the current simulation. Thus, for example, the system triggers increased restrictions if the weekly number of hospitalizations rises over the max threshold and distancing immediately becomes $\sigma_{\max}(a)$ which is age-variable: 70% of pre-pandemic levels in non-seniors and 50% in seniors. Then, once hospitalizations drop below the release threshold $T_{\min}$, 10% of the distancing is removed every $\tau$ weeks until reaching the minimum social distancing $\sigma_{\min}$. This value is not necessarily zero because we expect persistent features such as masking, work from home and avoidance of large social gatherings will continue to limit the number of interpersonal contacts relative to pre-pandemic levels.

*Vaccination mechanisms*. Original COVID-19 vaccine efficacy trials measured reductions in symptomatic disease. Therefore, it is unclear whether reductions in disease were mediated by totally prevented infections, or rather infections that were more likely to be asymptomatic. Therefore, we include the possibility of vaccines that work by several mechanisms and include three parameters controlling the effect of vaccination, $\mathrm{VE_{SUSC}}$, $\mathrm{VE_{SYMP}}$, $\mathrm{VE_{INF}}$. They are all vectors, so that if there are multiple vaccines defined, they can all have unique vaccine effectivenesses, likewise for any recovered classes. Vaccine effectivenesses are also strain-specific. The vaccine can completely block infection and reduce the number of vaccinated individuals that are susceptible by some fraction ($\mathrm{VE_{SUSC}}$), and thus modifies the left-hand side of the contact matrix, affecting susceptibility. Or, it can block symptomatic disease in individuals who are infected despite vaccination ($\mathrm{VE_{SYMP}}$), altering $p$ which controls the proportion of symptomatic and asymptomatic infections. Finally, it can decrease the possibility of onward transmission in individuals who are infected despite vaccination ($\mathrm{VE_{INF}}$), and thus it modifies the right-hand side of the contact matrix, altering $\beta^*$, thereby reducing transmission. Each vaccine efficacy ranges from 0-1. Furthermore, there is an additional vaccine eficacy, that against hospitalization (i.e. severe disease), conditional on being infected, $\mathrm{VE_H}$.

*Vaccination rollout*. The key parameters that govern vaccination distribution are the vaccination rate, $V_{rate}$, the vaccination distribution, $V_{dist}$, the vaccination coverage limit, $V_{coverage}$, and the vaccination priority, $V_{priority}$. The vaccination rate is the number of vaccines distributed per day and is set per vaccine. The vaccination distribution describes what percentage of vaccines are distributed to each age group. This can be set proportional to the percentage of the population for an equal distribution or can be used to prioritize certain age groups. For example, the inital vaccine rollout was targeted primarily at the elderly, then adults, and finally certain segments of the youngest age group. The vaccination coverage describes the upper limit of coverage to apply. It is applied per age group. The vacination priority parameter is a vector of age groups, and describes the order to reallocate available vaccines if one or more of the age groups has reached the coverage limit. All of these parameters can change throughout the vaccine rollout, i.e. to increase or decrease the doses available, using the standard temporal parameter framework.

Both susceptible and recovered individuals are eligible for vaccination, and are applied proportionally. While in theory individuals in the infected classes coul be vaccinated, this is not implemented in the model, and this follows advice to not be vaccinated while currenly ill with COVID. The V matrix describes which states are eligible for vaccination, as there can be a variable number of recovered states defined for the model, as well as what proportion of the total $V_{rate}$ doses available are for each vaccine.

**3. King County model calibration**.

*Data*. The model outputs were calibrated to three corresponding cumaltive metrics in King County WA: diagnosed cases, hospital admissions, and deaths. Thus corresponding to the cummlative values for the following states cases = $(\mathbf{DA1} + \mathbf{DA2} + \mathbf{DP} + \mathbf{DM} + \mathbf{DS} + \mathbf{DH})$, hosptializations = $(\mathbf{H} + \mathbf{DH})$, deaths = $(\mathbf{F} + \mathbf{DF})$. Each metric was tracked by age, which we consolidated into the 4 age groups $a$, so that calibration was performed against 12 metrics total.

Due to effects of weekends and weekdays, some noise in the data, and the tendancy of the daily time series to be auto-correlated, we took a 7 day smoothed average of 3 days before and after the day of interest, and then used weekly values of the metrics to calibrate against (that is 1 value every 7 days). Because the metrics were on very different scales, for example with many more cases than deaths or hospitalizations and differences across age classes, we normalized the metrics. For each of the 12 meteric time series, we divided by its mean, and applied the same procedure to the model output, also normaliing by the mean of the data metrics.

*Algorithm*. We used an Approximate Bayesian Computing (ABC) algorithm implemented in the EasyABC R package using the ABC_rejection function. This algorithm samples from the prior parameter distributions, runs the model to calculate the outputs, then calculates the distance between the model outputs and the data metrics. Because we already normalized the meterics, we used a simple Euclidian distance to select the 100 best fitting parameter sets. These formed the parameter posterior distributions, and we used this set of parameterizations in the simulations to incorporate parameter uncertainty.

*Calibrated parameters*. The model calibration was performed in two stages. An inital calibration was performed for the inital epidemic period, where the date of the inital case $t_0$ was a calibrated parameter, and the model was calibrated to data through April 30, 2020. The following parameters were calibrated: $\beta^*$, $\beta_D$, $t_0$, $\sigma(a)$, $\Delta_{IM}(a)$, $\Delta_{IS}(a)$, $\rho_A(a)$, $m(a)$. All parameters were given uniform prior distributions based on the best available data (either King County WA data or published studies). We obtained 100 parameterizations which we used for the posterior distributions of the parameters. We performed a second calibration for the epidemic state just before the simulation period, using data from April 1, 2021 through June 15, 2021. In order to retain the initial period parameterizations for $\beta^*$, $\beta_D$, and $m(a)$, we used the emprical posterior distribtion from the inital period calibration as the prior distribution for the final period c alibration. We performed the final period calibration with uniform priors for the following parameters: $\sigma(a)[1]$, $\sigma(a)[2]$, $\Delta_{IM}(a)$, $\Delta_{IS}(a)$, $\rho_A(a)$, where $\sigma(a)[1]$ was for the period April 1, 2021 through April 24, 2021, and $\sigma(a)[2]$ was for the period April 25, 2021 through June 15, 2021, in order to fit the increase and decline in cases during the calibration period. During both calibrations the $\sigma_{school}$ parameter was set to 1, reflecting the fact that schools were closed.

**4. Initial conditions**.

Cumulative incidence by age ($ci_a$) and current prevalence on June 1st, 2021 are back-calculated (see next section) from hospitalization in King County through July 31st, 2021 vaccination frequency by age group ($fv_a$) is derived from the CDC. Using this information, we populate all compartments using the following steps.

1. We subdivide the entire population, by age group ($N_a$) into three immune classes: "susceptible" ($S_a$), "vaccinated" ($V_a$), and"recovered" ($R_a$). This gives a total of 12 subpopulations.
2. The total prevalence by variant is set to the back calculated value for overall prevalence. It is assumed that, of initial infections, 80% are with the Alpha variant and 20% are with Delta. For each we create a 12x12 next generation matrix and use the normalized eigenvectors to subdivide the total prevalence by age and immune classes. This gives a total of 24 infected populations.
3. Each of the 24 infected populations is further subdivided into disease stages (such as symptomatic, diagnosed, hospitalized, etc.) using a quasi-steady state approximation.

All other initial populations, such as those corresponding to "waned" immune states, are assumed to be zero initially.

*Back-calculating infections* . Calculation of our initial conditions relies on back calculation of infections from hospitalization data, which we assume to be more consistently reported. We reconstruct infections by age, $\mathbf{x}_a$, from the combination of hospitalizations and non-hospitalized deaths by age, $\mathbf{y}_a$. We assume that the time from infection to hospitalization is exponentially distributed and furthermore we penalize large differences in $\mathbf{x}_a$ from one day to the next. We calculate $\mathbf{y}_a$ by solving the following regularized system with non-negative least squares.

$$(A^T A + \omega^2 \Gamma^T \Gamma)\mathbf{x}_a = A^T \mathbf{y}_a / IHR_a \quad (5)$$

The matrix $\Gamma$ is a first-order difference matrix and the matrix entry $A_{i\,j}$ represents the probability that an individual infected on day $j$ will become hospitalized on day $i$. $IHR$ is the infection hospitalization rate.

**5. References**

1. Davies, N. G. et al. Age-dependent effects in the transmission and control of COVID-19 epidemics. Nat. Med. 26, 1205–1211 (2020).

2. Abaluck, J. et al. The Impact of Community Masking on COVID 19A Cluster Randomized Trial in Bangladesh. https://www.poverty-action.org/publication/impact-community-masking-covid-19-cluster-randomized-trial-bangladesh (2021).

3. Bracis, C. et al. Widespread testing, case isolation and contact tracing may allow safe school reopening with continued moderate physical distancing: A modeling analysis of King County, WA data. Infect. Dis. Model. 6, 24–35 (2021).

4. Sheikh, A., McMenamin, J., Taylor, B. and Robertson, C. SARS-CoV-2 Delta VOC in Scotland: demographics, risk of hospital admission, and vaccine effectiveness. Lancet 397, 2461–2462 (2021).

5. Bernal, J. L. et al. Effectiveness of COVID-19 vaccines against the B . 1 . 617 . 2 variant. medRxiv https://www.medrxiv.org/content/10.1101/2021.05.22.21257658v1.full.pdf (2021) doi:https://doi.org/10.1101/2021.05.22.21257658;

6. Davies, N. G. et al. Estimated transmissibility and impact of SARS-CoV-2 lineage B.1.1.7 in England. Science (80-. ). 372, eabg3055 (2021).

7. Volz, E. et al. Assessing transmissibility of SARS-CoV-2 lineage B.1.1.7 in England. Nature 593, 266–269 (2021).

8. Dhar, A. M. S., Marwal, R., Vs, R. and Ponnusamy, K. Genomic characterization and Epidemiology of an emerging SARS CoV-2 variant in Delhi India medRxiv 1 - 17 (2021) doi.org10.1101/2021.06.02.21258076

9. Public Health England. SARS-CoV-2 variants of concern and variants under investigation in England. (2021).

10. Fisman, D. and Tuite, A. Progressive Increase in Virulence of Novel SARS-CoV-2 Variants in Ontario, Canada. medRxiv vol. 6 (2021).

11. Dagpunar, J. Interim estimates of increased transmissibility, growth rate, and reproduction number of the Covid-19 B.1.617.2 variant of concern in the United Kingdom. medRxiv (2021) doi:https://doi.org/10.1101/2021.06.03.21258293.

12. Ong, S. W. X. et al. Clinical and virological features of SARS-CoV-2 variants of concern: a retrospective cohort study comparing B.1.1.7 (Alpha), B.1.315 (Beta), and B.1.617.2 (Delta). Clin. Infect. Dis. (2021) doi:https://doi.org/10.1093/cid/ciab721.

13. He, X. et al. Temporal dynamics in viral shedding and transmissibility of COVID-19. Nat. Med. 26, 672–675 (2020).

14. Walsh, K. A. et al. The duration of infectiousness of individuals infected with SARS-CoV-2. J. Infect. 81, 847–856 (2020).

15. Havers, F. P. et al. Seroprevalence of Antibodies to SARS-CoV-2 in 10 Sites in the United States, March 23-May 12, 2020. JAMA Intern. Med. 180, 1776–1786 (2020).

16. Gray, W. K. et al. Variability in COVID-19 in-hospital mortality rates between national health service trusts and regions in England: A national observational study for the Getting It Right First Time Programme. EClinicalMedicine 35, 100859 (2021).

17. Davies, N. G. et al. Age-dependent effects in the transmission and control of COVID-19 epidemics. Nat. Med. 26, 1205–1211 (2020).

18. Centers for Disease Control and Prevention, Interim Public Health Recommendations for Fully Vaccinated People https://www.cdc.gov/coronavirus/2019-ncov/vaccines/fully-vaccinated-guidance.html#anchor_1619526549276 (accessed 9.21.21).

19. King County, Summary of COVID-19 vaccination among King County residents https://kingcounty.gov/depts/health/covid-19/data/vaccination.aspx (accessed 9.21.21).

20. Washington Governor Jay Inslee, Inslee signs emergency proclamation requiring in-person education opportunities for public K-12 schools. https://www.governor.wa.gov/news-media/inslee-signs-emergency-proclamation-requiring-person-education-opportunities-public-k-12 (accessed 9.27.21).

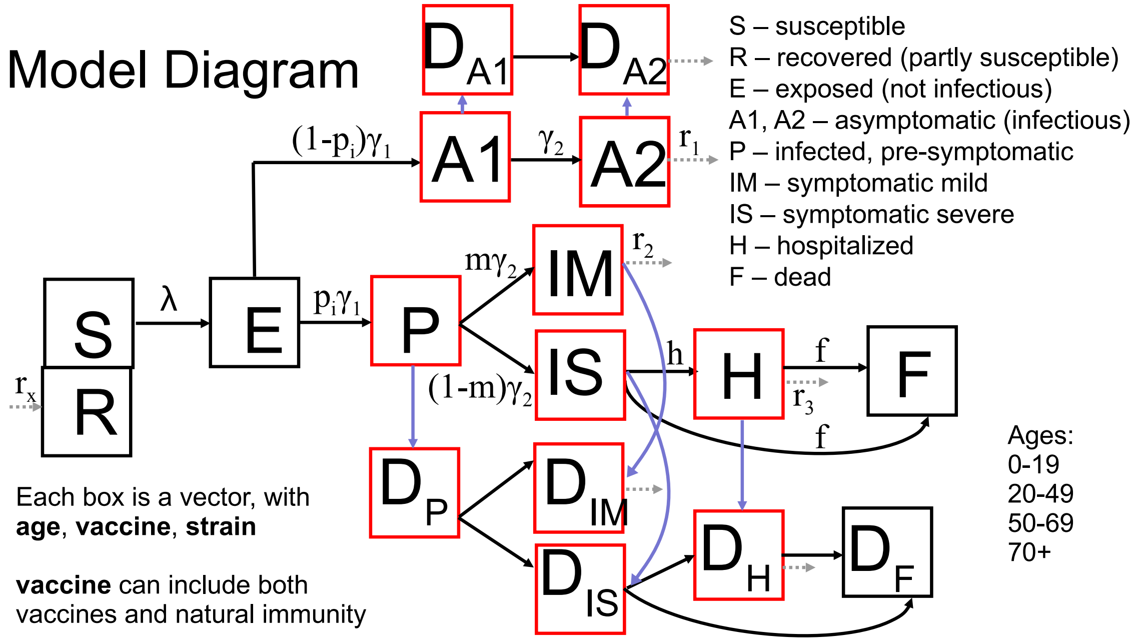


**Fig S1.** Model diagram.

**Supplementary Table 2.** Table of model parameters, which can depend on the variables age $a$, vaccination status $v$, and strain $q$. Note that the model has the flexibility to make any variable time varying.

| Parameter | Description | Values | Reference |
|---|---|---|---|
| $\lambda$ | Force of infection | See equation | Estimated |
| $\kappa$ | Relative susceptibility by age | 0.5, 1, 1, 1 | [1] |
| $\sigma_{min}$ | Minimum level of social distancing by age | 0.1,0.1,0.1,0.2 | [2] |
| $\sigma_{max}$ | Maximum level of social distancing by age | 0.3,0.3,0.3,0.5 | [3] |
| $\sigma_{inc}$ | Increment when releasing dynamic social distancing | 0.10 | |
| $\tau$ | Time period over which to calculate threshold metric - days | 7 | |
| $\mathrm{VE}_{\mathrm{SUSC}}$ | Vaccine efficacy - susceptibility- alpha, delta | 0.91,0.82 | [4, 5] |
| $\mathrm{VE}_{\mathrm{SYMP}}$ | Vaccine efficacy - symptoms - alpha, delta | 0.34, 0.34 | [4] |
| $\mathrm{VE}_{\mathrm{INF}}$ | Vaccine efficacy - transmission | 0 | Assumed |
| $\mathrm{VE}_{\mathrm{H}}$ | Vaccine efficacy - hospitalization - alpha, delta | 0.67,0.67 | [4] |
| $\nu_{inf}(q)$ | Relative transmissibility by strain - original, alpha, delta | 1, 1.5, 2.4 | [6-9] |
| $\nu_{sev}(q)$ | Relative severity by strain - original, alpha, delta | 1, 1.5, 1.5 | [10-12] |
| $id$ | Duration of symptomatic infectiousness (days) | 5 | [13] |
| $t_0$ | Day of first infection | 7-25 | Calibrated |
| $\gamma_1$ | Waiting time for exposed | 0.33 | [13] |
| $\gamma_2$ | Waiting time for pre-symptomatic (P or A1) | 0.5 | [13] |
| $r_A$ | Recovery rate for asymptomatic | 0.2 | |
| $r_M$ | Recovery rate for mild symptomatic | $1/id$ | [13] |
| $r_H$ | Recovery rate for hospitalized (severe symptomatic) | 1/15 | [14] |
| $\rho_A(a)$ | Relative likelihood of asymptomatic to be diagnosed w.r.t. symptomatic | 0.15-0.3, 0.15-0.3, 0.15-0.3, 0.15-0.3 | Calibrated |
| $\Delta_{IM}(a)$ | Diagnosis rate for mild infections | 0.15-0.35, 0.15-0.35, 0.15-0.35, 0.15-0.35 | Calibrated, [15] |
| $\Delta_{IS}(a)$ | Diagnosis rate for severe infections - by age | 0.2-0.5, 0.2-0.5, 0.2-0.5, 0.2-0.5 | Calibrated |
| $\Delta_H$ | Diagnosis rate for hospitalized | 0.75 | Estimated |
| $h(a)$ | Hospitalization rate | 0.15 | Estimated |
| $hfr$ | Hospital fatality ratio | 0.009, 0.024, 0.105, 0.318 | Estimated, [16] |
| $inf_pre$ | Proportion of infections due to pre-symptomatic individuals | 0.44 | [13] |
| $\pi$ | Proportion symptomatic - by age | 0.25, 0.33, 0.55, 0.70 | [17] |
| $m$ | Proportion of symptomatic infections which are mild - by age | 0.988-0.996, 0.96-0.995, 0.87-0.96, 0.62-0.87 | Calibrated |
| $\beta*$ | Base transmission rate | 0.01-0.04 | Calibrated |
| $\beta_{A1}$ | Relative transmission rate for A1 | $0.75 * \beta_P$ | |
| $\beta_{A2}$ | Relative transmission rate for A2 | 0.75 | |
| $\beta_P$ | Relative transmission rate for P | $inf_{pre}/(1 - inf_{pre}) * (id * \gamma_2)$, | [13] |
| $\beta_M$ | Relative transmission rate for IM | 1 | Assumed |
| $\beta_S$ | Relative transmission rate for IS | 1 | Assumed |
| $\beta_H$ | Relative transmission rate for H | 0 | Assumed |
| $\beta_D$ | Reduction in transmission due to being diagnosed | 0.25-0.9 | Calibrated |
| $V_{rate}$ | Vaccine doses per day | 5000 | Estimated |
| $V_{dist}$ | Age distribution of vaccines | 0.2293, 0.4552, 0.2350, 0.0805 | Estimated |
| $V_{coverage}$ | Upper coverage limit of vaccination | 0.8, 0.85, 0.9 | Scenarios |
| $V_{priority}$ | Age group priority order for vaccination | 4:1 | Assumed |

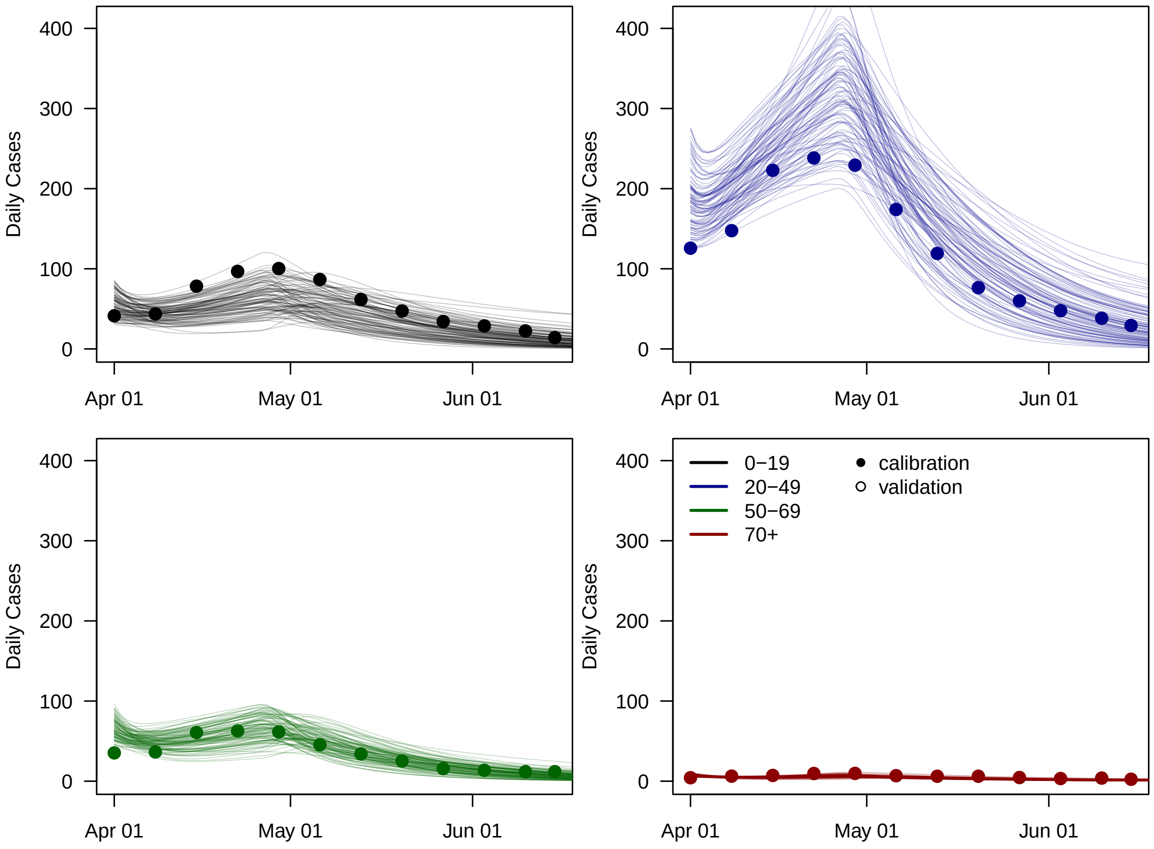


**Fig S2.** The 100 parameterizations shown against data for diagnosed cases (points) by age group for the calibration period.

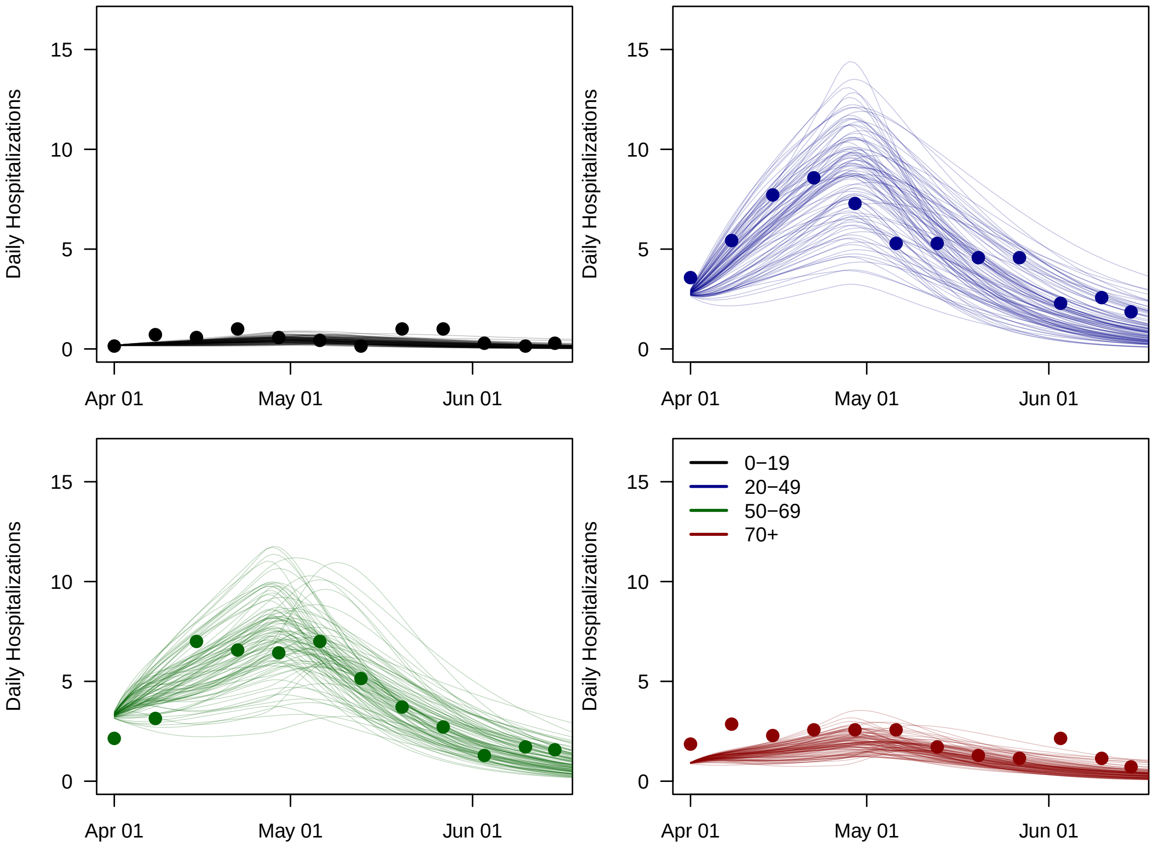


**Fig S3.** The 100 parameterizations shown against data for hospitalizations (points) by age group for the calibration period.

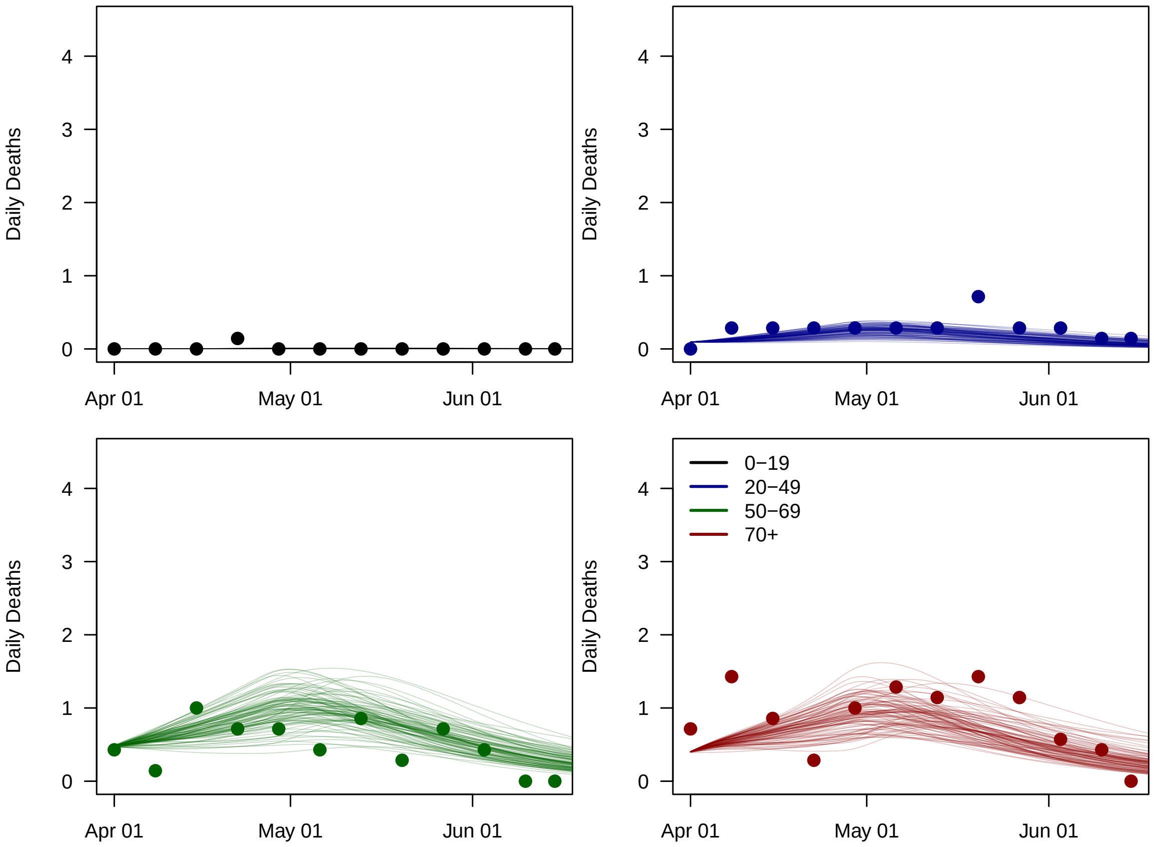


**Fig S4.** The 100 parameterizations shown against data for deaths (points) by age group for the calibration period.

**5. Additional results**

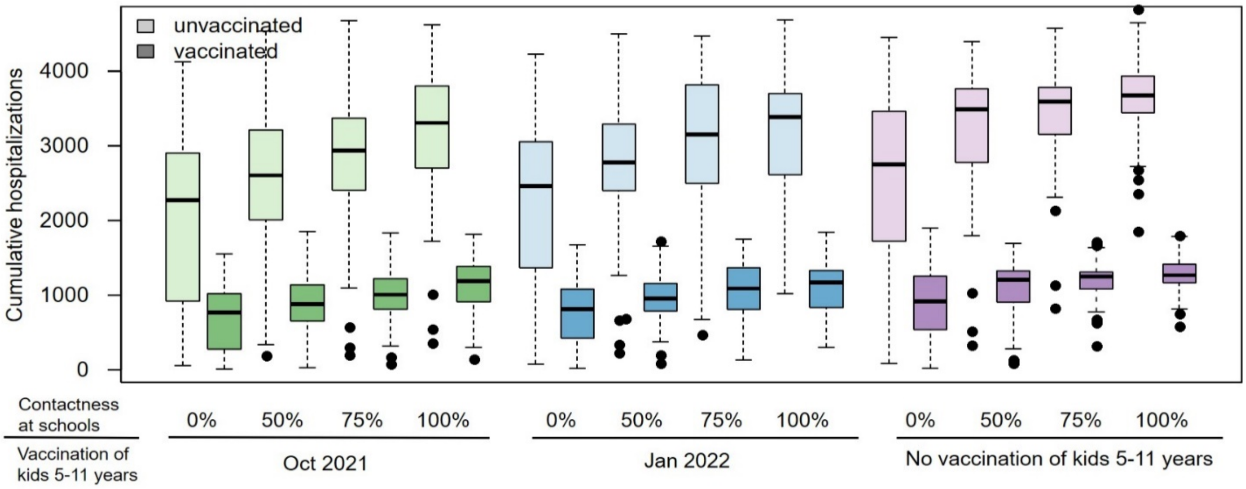


**Fig S5. Cumulative hospitalizations by vaccination status**

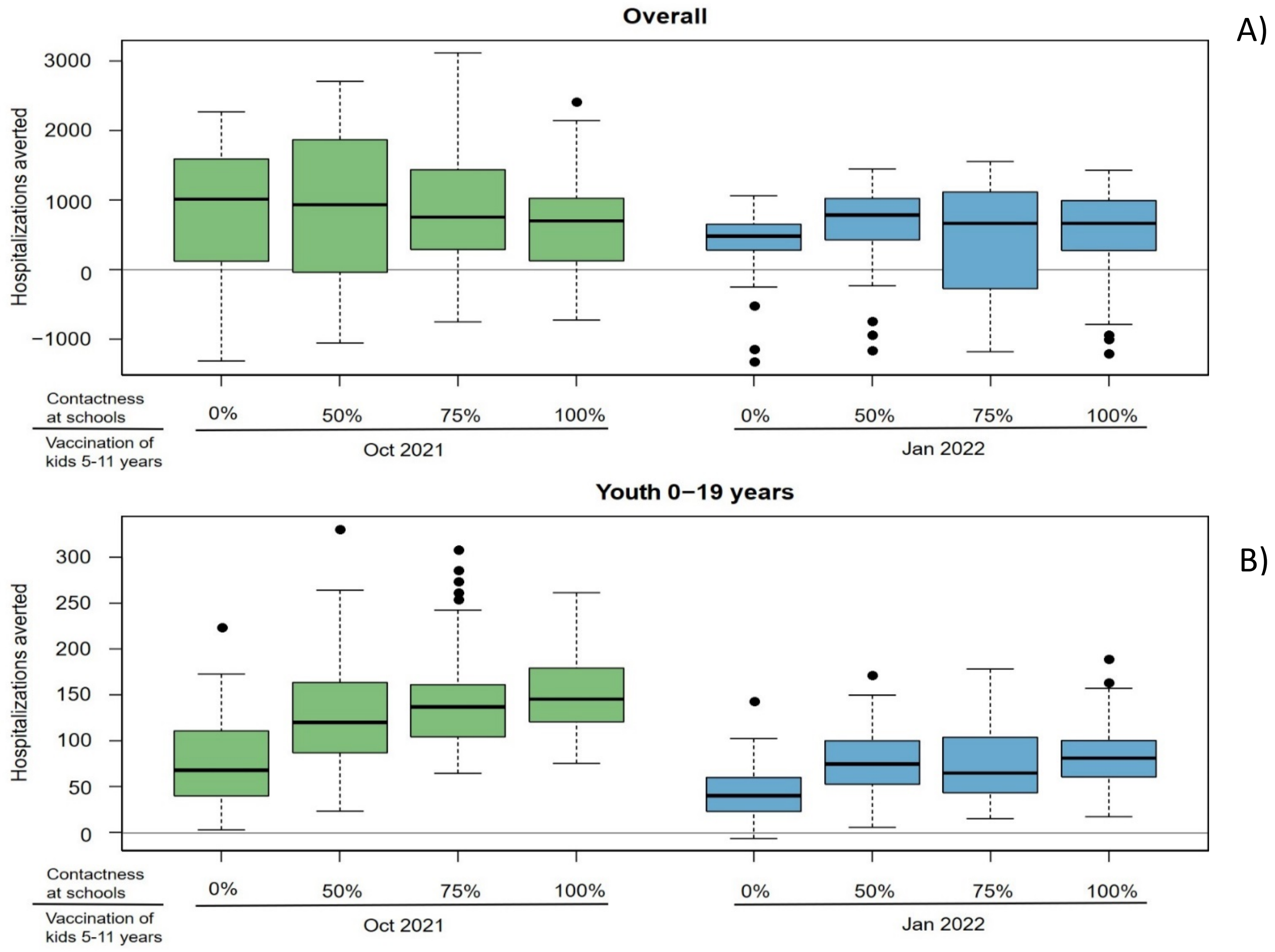


**Fig S6. Reduction in hospitalizations due to expanding vaccination to kids 5-11 years:** ***A)*** *Overall;* ***B)*** *Among the youngest group (0-19 years old) only. Boxplots show the distribution of the absolute difference between simulations with different start of vaccinating kids 5-11 years compared to simulations without vaccinating this age group. Negative differences are results of the reactive social distancing rules imposed.*

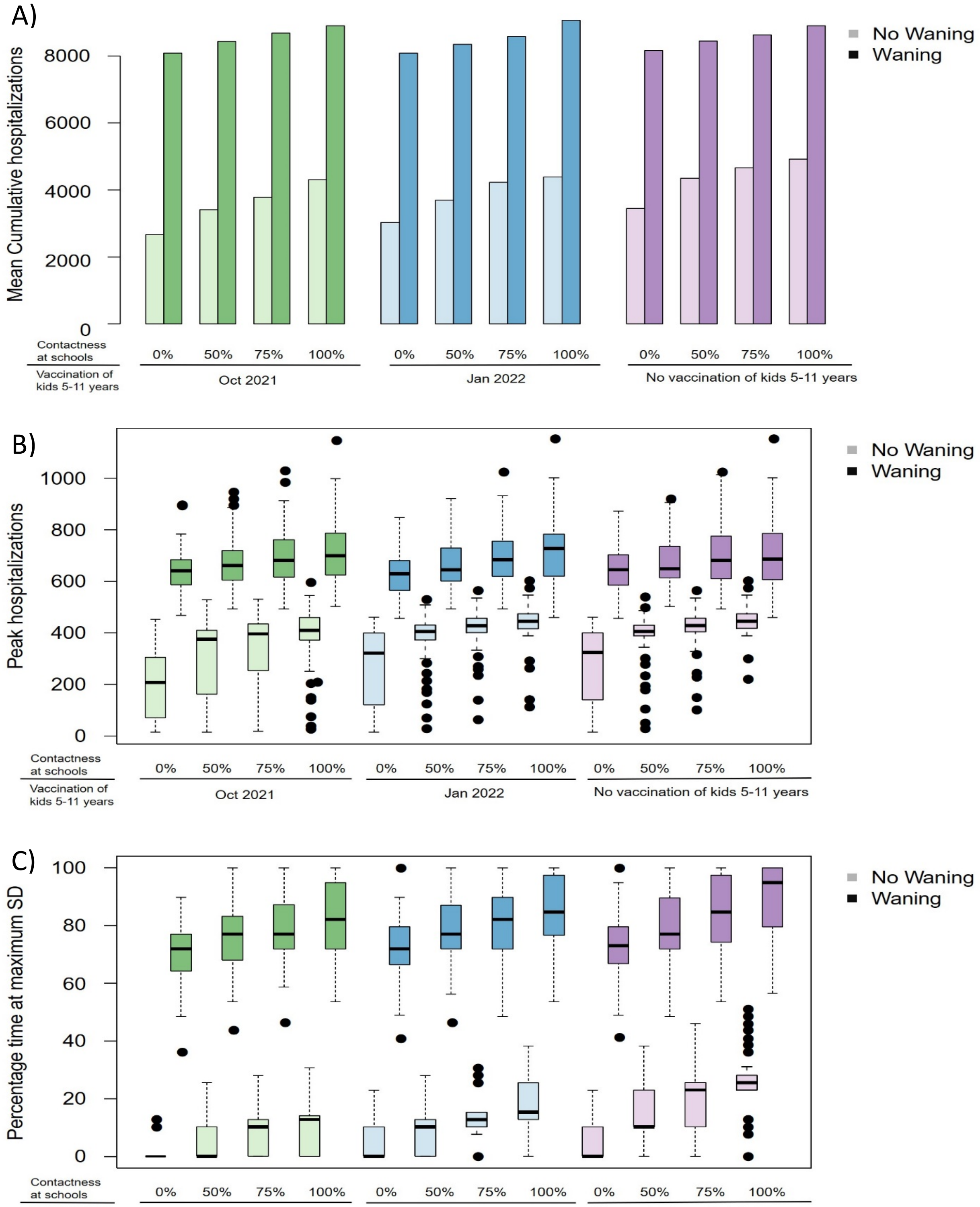


**Fig S7. Effects of efficacy waning.** ***A)*** *Mean cumulative hospitalizations;* ***B)*** *Maximum number of hospitalized people over the school year and* ***C)*** *Percentage time of the school year under maximum restricted social distancing (SD) under scenarios assuming waning protection from vaccination and prior infections compared to scenarios without waning.*
